# Expanded chromosomal microarray comprising screening for spinal muscular atrophy and monogenic diseases3

**DOI:** 10.1101/2024.11.19.24309471

**Authors:** Xiaorui Luan, Naixin Xu, Yaojun Xie, Weihui Shi, Xianling Cao, Xuanyou Zhou, Songchang Chen, Chenming Xu

**Author notes:** Corresponding author: Songchang Chen, Chenming Xu. These authors contributed equally to this work (Xiaorui Luan, Naixin Xu). **Date sharing:** All the data generated or analyzed in this study are included in this published article. **Ethical approval**: Ethical approval for this study was “Construction and Application Evaluation of a Prenatal Detection System Based on Novel Microarray Technology” obtained from ethical Committee of the Obstetrics & Gynecology Hospital of Fudan University (2022-46). **Patient consent:** Informed consent was obtained from all the study participants at the time of providing samples.

## Abstract

**Background:** Copy number variants platforms, as critical supports for genetic diagnosis, have been well implemented in prenatal diagnosis. However, numerous severe conditions with underlying single-gene defects are not included in current invasive prenatal screening. To bridge this gap, an expanded chromosomal microarray analysis was developed, employing a meticulous designed single nucleotide polymorphism chip. This chip incorporated additional probes to augment its efficacy in screening for spinal muscular atrophy and diagnosing monogenic disorders.

**Objective(s):** This study aimed to evaluate the accuracy, efficacy, and incremental yield of expanded chromosomal microarray, compared with karyotype analysis and low-depth genome sequencing for routine prenatal diagnosis.

**Study Design:** In this prospective study, total of 512 fetuses were included in this study. In this study three distinct diagnostic techniques-karyotype analysis, low-depth genome sequencing, and expanded chromosomal microarray-were processed to evaluate each sample. Aneuploidies and multigene copy number variations were detected and analyzed in a blinded fashion. *SMN1* exonic copy number variations were confirmed by multiplex ligation-dependent probe amplification and single nucleotide variations were confirmed by sanger sequencing.

**Results:** Overall, expanded chromosomal microarray identified genetic abnormalities in 91 out of 512 cases (17.6%). The encountered rate was significantly higher than the rates observed with low-depth genome sequencing (66 out of 512 cases, 12.9%) and conventional chromosome karyotyping (42 out of 512 cases, 8.2%). Expanded chromosomal microarray not only detected all these non-mosaic aneuploidies and copy number variations in 62(12.1%) diagnosed cases identified by low-depth genome sequencing (low-depth genome sequencing), but also detected 9 cases with regions of homozygosity, 10(2.0%) cases with exonic deletions (*SMN1* and *DMD*), and 13(2.5%) cases with single nucleotide variations.

**Conclusions:** Compared with low-depth GS, expanded chromosomal microarray increased the additional detection rate by 4.7% (24/512). Compared with traditional chromosomal microarray, expanded chromosomal microarray increased the additional detection rate by 3.9% (20/512) in 512 fetuses. Although the expanded chromosomal microarray (ECMA) has limited accuracy for detecting single nucleotide variations, its screening capacity is significantly enhanced when complemented with Sanger sequencing validation. Using expanded chromosomal microarray, we detected not only copy number variations, but also exonic deletions, regions of homozygosity with high accuracy in an acceptable turnaround time (2-3 weeks). Our results suggest that expanded chromosomal microarray has the potential to be a promising prenatal diagnostic tool with incremental yield of screening exonic copy number variations in *SMN1*.

## Introduction

Fetal structural abnormalities occur in approximately 3% of pregnancies^1^. In the current landscape of prenatal diagnosis, both karyotype analysis, low-depth genome sequencing (low-depth GS) and chromosomal microarray analysis (CMA) have been widely adopted as standard techniques for invasive prenatal diagnostic procedures^2,3^. Among these techniques, CMA has been recommended to any patient choosing to undergo invasive diagnostic testing^4^. CMA has facilitated the identification of chromosomal aneuploidies and multigene copy number variations (CNVs) in prenatal settings and also permitted the detection of submicroscopic genomic imbalances across the entire genome, thereby enhancing the diagnostic yield in genetic studies^4–6^.

Prenatal genetic testing aids in optimizing neonatal outcomes by providing necessary care for affected infants^7–9^. Among these, CNVs and single nucleotide variants (SNVs) play a critical role in the etiology of numerous genetic disorders^10,11^. Carrier screening for exonic deletion disorders such as spinal muscular atrophy (SMA) and Duchenne muscular dystrophy (DMD) is essential^12,13^. As a fatal genetic disease that leads to severe muscle weakness, SMA affects about 1 in 12,000 people worldwide^14,15^. The molecular diagnosis of the SMA patients and carriers consists of the detection of the absence of exon 7 of the *SMN1* gene^16^. Notably, the preemptive diagnosis and intervention with disease-modifying treatment in neonates prior to symptom onset demonstrates superior outcomes compared to post-symptomatic administration in SMA management^15,16^. DMD, an X-linked recessive genetic condition manifesting in progressive muscle wasting and weakness, stands as the predominant form of childhood muscular dystrophy^17^. DMD is the most common form of muscular dystrophy, with a global incidence rate of 19.8 cases per 100,000 live male births^18^. The underlying genetics dictate a predilection for male affliction, whereas female carriers often remain asymptomatic or exhibit mild symptoms that can be easily overlooked. The genetic diagnosis of the 65%-85% DMD patients is the duplication or deletion of the DMD gene^19^. To date, the absence of targeted and efficacious pharmacological interventions remains a significant challenge in the clinical management of DMD^19–21^. The detection of SMA and DMD relies on multiplex ligation-dependent probe amplification (MLPA) technology. Although MLPA has high accuracy, it requires a substantial amount of sample. This renders it impractical for widespread use as a screening tool for precious invasive prenatal diagnostic samples. Besides, over half of pediatric and fetal cases with single-gene defects are ascribed to monogenic variants^22–24^. The single nucleotide polymorphism array (SNP array) has been widely employed in the detection of variants^25,26^. The successful deployment of SNP array for a given application is critically dependent on array design and appropriate extensive experimental validation^10^. In recent study, a SNP array targeting not only aneuploidies, multigene CNVs, but also SMA carrier screening and SNVs has been developed. However, the accuracy of such a comprehensive diagnose approach combined with screening function has not yet been explored in routine clinical settings through prospective cohort studies.

This study aims to evaluate the detection capabilities of expanded chromosomal microarray (ECMA) analysis which targets three of the most prevalent types of pathogenic genetic variants: aneuploidies, CNVs and monogenic variants. Additionally, this prospective study explores the ECMA platform’s capability in identifying multigene CNVs and its utility in exonic CNV carrier screening for conditions such as SMA and DMD. The investigation also encompasses an analysis of the clinical characteristics of the participants, providing a comprehensive overview of the genetic testing indications and the demographic profile of the study population.

## Materials and Methods

### Subjects

A flowchart of the study is presented in Figure 1. Between March 2022 and July 2023, 498 pregnant women subjected to invasive prenatal diagnostic procedures were enrolled, among whom 13 had twin pregnancies. All fetal specimens were collected by amniocentesis or chorionic villus sampling. All cases were investigated using Low-depth GS as a routine test for detecting CNVs, using karyotyping for chromosomal analysis. In parallel, the remaining deoxyribonucleic acid (DNA) was used for ECMA and validation. Ethical approval for this study was “Construction and Application Evaluation of a Prenatal Detection System Based on Novel Microarray Technology” obtained from ethical Committee of The Obstetrics & Gynecology Hospital of Fudan University (2022-46).

**Figure 1.**
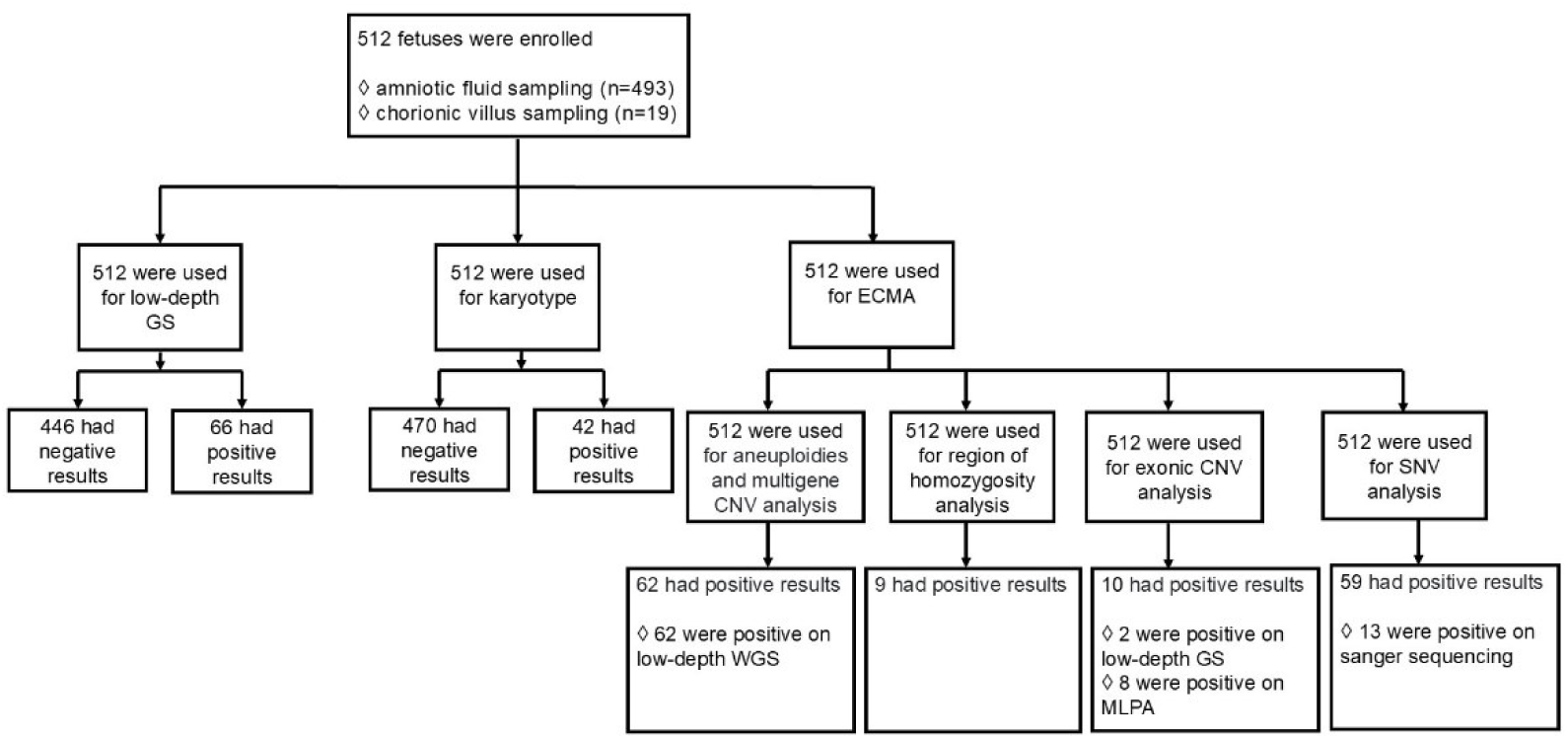
Flowchart of the study. A total of 512 fetuses were enrolled. Among them, 493 were collected by amniotic fluid, 19 were collected by chorionic villus. Three genetic testing methods applied to the prenatal samples: Karyotype analysis (n=512), low depth genome sequencing (GS)(n=512), and expanded chromosome microarray (ECMA)(n=512). For validation of the findings, two techniques were employed: Sanger sequencing (applied to n=110 cases) and multiplex ligation-dependent probe amplification (MLPA) (performed on n=8 cases).

### ECMA

Genomic DNA was extracted from 10 ml amniotic fluid samples or 5 mg chorionic villus samples using a QIAamp DNA Mini Kit (Qiagen, Germany). CMA platforms CNVPlus were introduced in this study. CNVPlus platform designed based on Gene Titan (Thermo Fisher Scientific, USA) includes scan instrument, analyze platform, microarray. The probes of microarray including 620,000 CNV probes and 350,000 SNV probes. *SMN1* and *DMD* are targets dense coverage area. The method for interpreting SNVs and SMA screening results via ECMA is illustrated in Figure S1. A total of 50–100 ng fetal DNA was tested. The procedures were performed according to the method described by Ronald J. Wapner^27^. Results are shown in Table S1.

### Variants Classification

CNVs and SNVs were classified according to the criteria established by The American College of Medical Genetics and Genomics (ACMG) and the Clinical Genome Resource (ClinGen), utilizing the tool named Franklin, which is available at https://franklin.genoox.com/clinical-db/home^28,29^.

### Screening gene list

In accordance with the genetic screening criteria for both prenatal and preconception periods, secondary findings identified through the whole genome sequencing recommended by ACMG were considered. This process included the list of diseases for newborns and prenatal screening in China. A total of 98 genes were meticulously selected for inclusion in the study, with probe designs specifically tailored to target these genes on the microarray chip. (Table S2)^30–33^.

### Karyotype analysis, Low-depth GS and MLPA

The karyotype procedures were performed according to the method described by Ronald J. Wapner^27^. Amniocentesis was performed by standard procedures. Amniocyte DNA was analyzed by CNV-Seq (Berry) China (with a chromosome resolution of 0.1 Mb. The procedures were performed according to the method described by Wang^34^. A total of 100-200 ng fetal DNA was tested with MLPA Probemix, as described before (P060-100R)^35^.

## Results

### Clinical characteristics of the participants

In the present investigation, a total of 498 pregnant women, among whom 13 had twin pregnancies, were subjected to invasive prenatal diagnostic procedures and subsequently enrolled in the study. The mean maternal age was 32.7 years (range, 17-46 years). The mean age of fetuses was 19.7 weeks (range,12-33 weeks). (see Table 1).

**Table 1.**
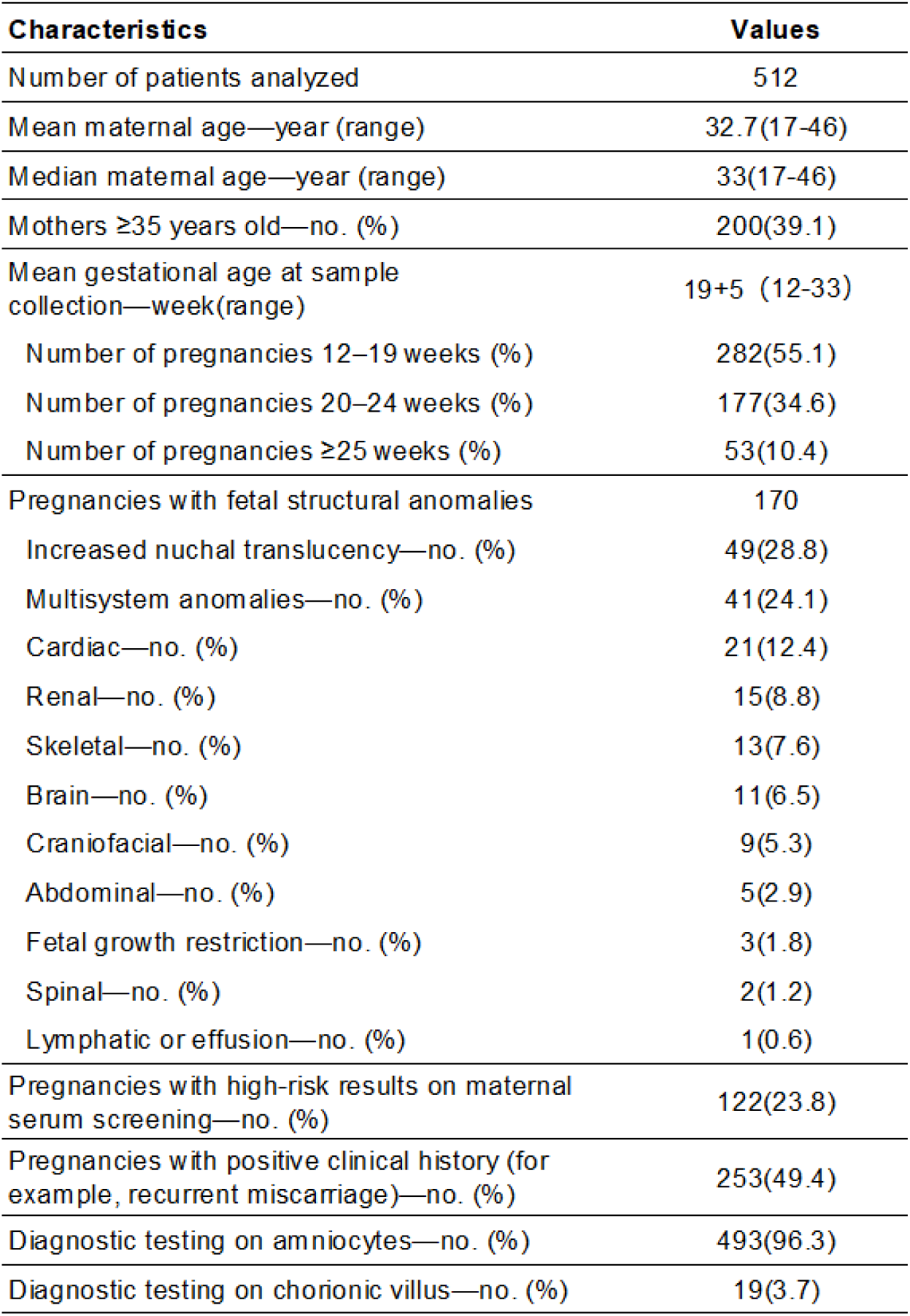
Demographic and clinical characteristics.

Structural anomalies identified by ultrasonography (n=170), chromosome abnormalities identified by maternal serum screening (n=122), recurrent miscarriage (≥2) (n=224), advanced maternal age (≥35) (n=200), validation of clinical preimplantation genetic testing (PGT) (n=34), parental carrier of a genetic disorder (n=32), previous fetus or child with autosomal trisomy or sex chromosome aneuploidy (n=15), prior child with structural birth defect (n=14). 19 participants were identified who did not meet the inclusion criteria for any of the previously defined groups. (see Table 1).

### CNV results of ECMA

#### aneuploidies

In this study, non-mosaic aneuploidies were assessed using three distinct aneuploidies diagnostic techniques-karyotype analysis, Low-depth GS, and ECMA-to evaluate a cohort of samples (see Table 2). Upon detailed examination of the non-mosaic results, it was observed that all three methodologies displayed identical sensitivity for Trisomy 21, detecting 18 cases apiece. Consistently, the number of non-mosaic Trisomy 18 cases detected was also uniform, being 3 for each method. Furthermore, the diagnoses of Klinefelter Syndrome, Trisomy X, Turner Syndrome and Jacobs Syndrome showed perfect agreement, with 3, 2, 2, 1 positive instance identified through all platforms. Consequently, this result supports the notion that these technologies can be considered equally effective in the context of detecting non-mosaic aneuploidies (see Table S1, S3).

**Table 2.** Comparative information of 512 fetuses detected by ECMA, low-depth GS, and karyotype analysis.

| Diagnostic yield | Karyotype analysis | low-depth GS | ECMA |
| --- | --- | --- | --- |
| Aneuploidies (non-mosaic) | 31/512 | 31/512 | 31/512 |
| Aneuploidies (mosaic) | 6/512 | 4/512 | 2/512 |
| Multigene CNVs | 5/512 | 29/512 | 29/512 |
| Regions of homozygosity | - | - | 9/512 |
| SNVs | - | - | 13/512 |
| Exonic CNVs | - | 2/505 | 10/512 |
| **Total** | **42/512**<br>**8.2%** | **66/512**<br>**12.9%** | **91/512 \***<br>17.7% |

\*: 2 type positive results on 3 samples (Sample ID: 341, 508, 509). ECMA: expanded chromosomal microarray, low-depth GS: low-depth genome sequencing, CNV: copy number variation.

The present investigation identified 2 mosaic chromosome abnormalities, confirmed by multiple diagnostic modalities including ECMA, karyotyping, and Low-depth GS. One such instance illustrated a 20% mosaic trisomy of chromosome 7, while the other case characterized by a mosaic ratio of 50% for the 45,XO constitution.

Upon comparative analysis, it was observed that ECMA technique missed the identification of mosaic anomalies in 2 fetal amniotic fluid samples where the proportion of mosaicism was lower than 20%. These cases entailed a specimen with a 10% mosaic trisomy 21 (Sample ID 28), along with a case (Sample ID 60) of mosaic Turner syndrome (45,XO) featuring mosaic ratios of 14% (see Table S1).

#### Multigene CNV

In the current study, using the ECMA platform, we identified 29 copy number variations (CNVs) (see Table S1, S4). We observed that none of CNVs shared the same genomic localization. Specifically, two distinct samples were found to contain two independent CNV fragments each, and one single sample manifested the presence of three discrete CNV regions. These CNVs, each encompassing an array of genes, are distributed across 15 distinct autosomes and the sex chromosomes. The dimensions of these CNVs show a considerable range, with the smallest identified CNV measuring 0.14 Mb and the largest reaching 4.8 Mb.

The ECMA technology offers a more extensive probe coverage on the Y chromosome. Compared to Low-depth GS covering chrY:6430000-23750000, ECMA covering chrY:2654333-28793765. ECMA facilitated the detection of a breakpoint between chrY:23883894-23955828 (hg19) in case 261, whereas Low-depth GS analysis failed to detect the breakpoint. By integrating the results from ECMA, Fluorescence In Situ Hybridization (FISH), and karyotyping, it is possible to determine that the actual chromosomal constitution of the sample may be 46,X,idic (Y)(q11.2) (see Figure 2).

**Figure 2.**
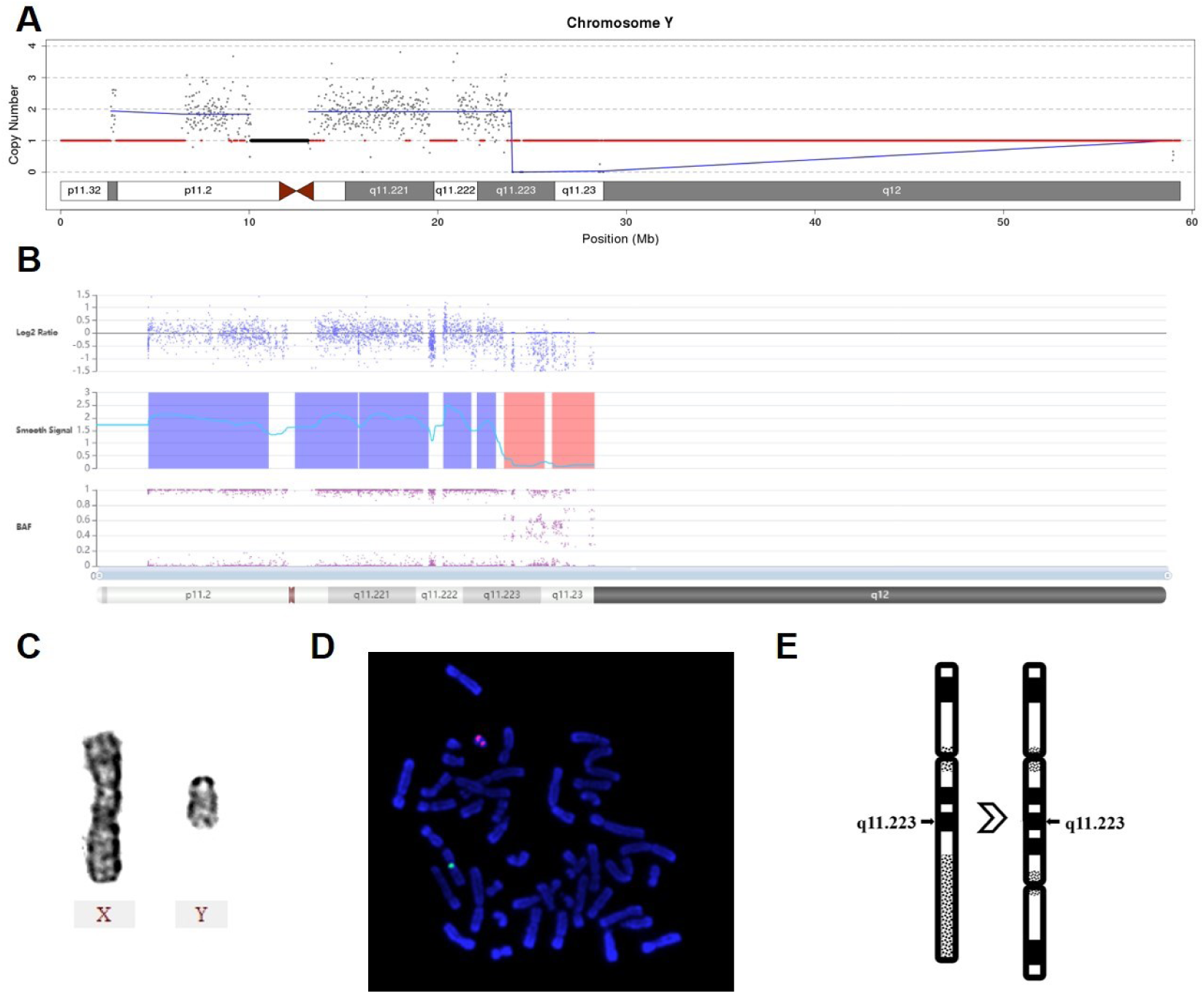
Identification of isodicentric chromosome breakpoint on chromosome Y. Low depth genome sequencing result (A) and expanded chromosome microarray result (B) of the chromosome Y. (C) Karyotype analysis results of sex chromosome. (D) Fluorescence in situ hybridization results. Green fluorescence probe is DXZ1 located in Xp11.1-q11.1, red fluorescence probe is DYZ3 located in Yp11.1-q11.1. (E) Diagram of results 46,X,idic (Y)(q11.2).

#### Exonic CNV

Molecular genetic analysis identified a homozygous deletion of exon 7 in the SMN1 with positive results confirms the diagnosis of spinal muscular atrophy (SMA), while individuals with a heterozygous deletion of exon 7 in the SMN1 gene were definitively identified as carriers of SMA.

In this study, a total of eight individuals with heterozygous deletions of the SMN1 gene were identified by ECMA as carriers of SMA (Table 3, S1).

**Table 3.**
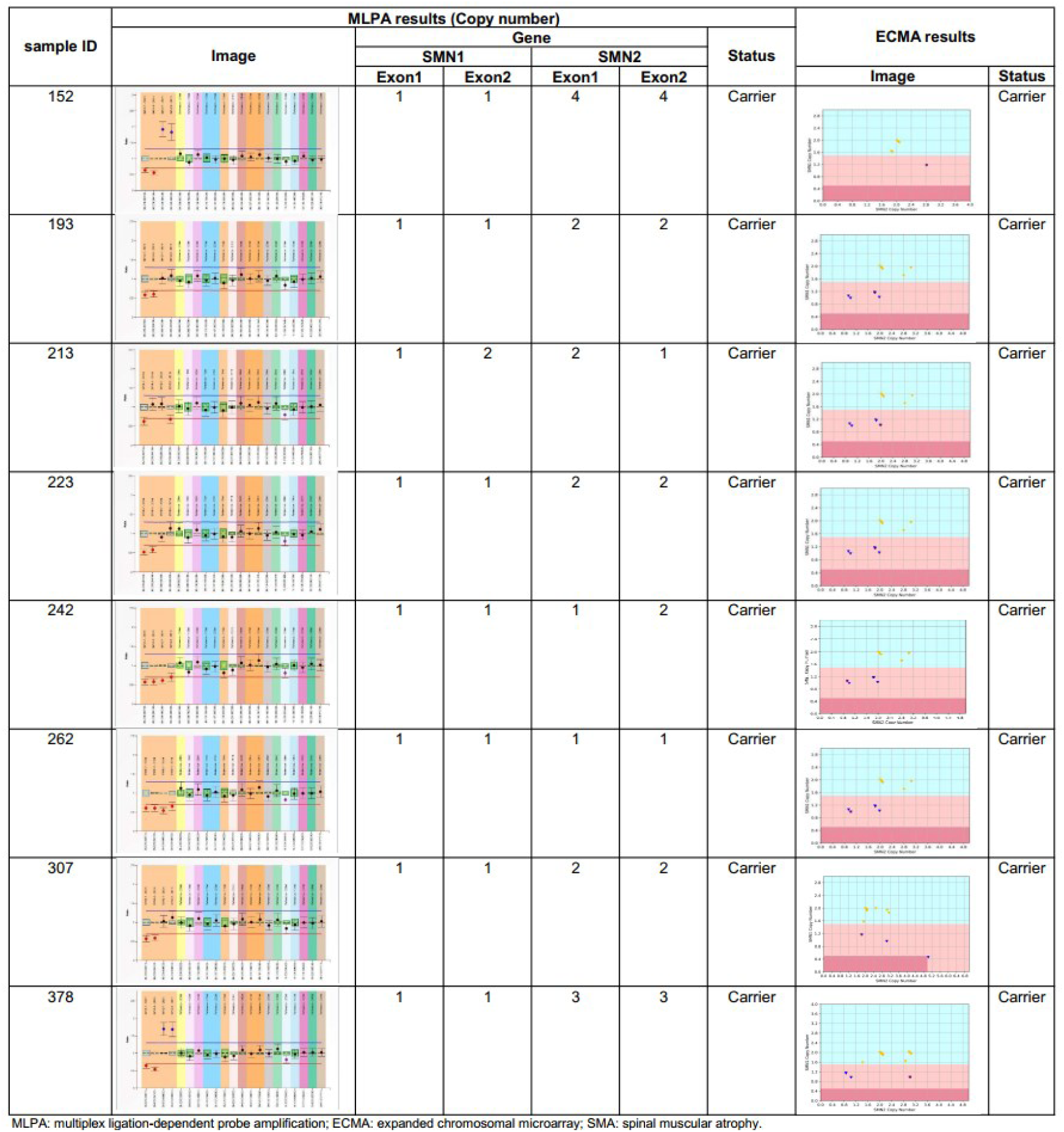
8 SMA carrier screening results of ECMA and MLPA.

One was found to have a heterozygous deletion of Exon 7 in the SMN1 gene and Exon 8 in the SMN2 gene, with normal copy numbers for Exon 8 in the SMN1 gene and Exon 7 in the SMN2 gene (Case 378). The other seven individuals exhibited heterozygous deletions of Exon 7 and Exon 8 in the SMN1 gene. Among them, one individual displayed a copy number of 1 for both Exon 7 and Exon 8 in the SMN2 gene (Case 262), while three individuals had no abnormalities in their SMN2 copy numbers (Case 193, 223, 307). Two individuals presented with an increased copy number for Exon 7 and Exon 8 in the SMN2 gene (Case 196, 378), and one individual had a copy number of 1 for Exon 7 and a normal copy number for Exon 8 in the SMN2 gene (Case 213) (Table 3).

Utilizing the MLPA methodology for stringent confirmation, it was ascertained that all subjects exhibiting heterozygous deletion of the SMN1 gene as detected by the ECMA platform indeed represent genuine positive cases.

Duchenne muscular dystrophy (DMD) is a X-linked disease caused by frameshifting mutations or nonsense mutations in the DMD gene, leading to complete absence of dystrophin and progressive degeneration of skeletal musculature and myocardium^19^. Our screening revealed multi-exon deletions in two samples: a deletion of exons 3–7 in Case 382 and a deletion of exons 45–55 in Case 383. (see Table S5)

#### SNV results

In the course of this study, 111 single nucleotide variants (SNVs) loci were preliminarily flagged as potential mutations following an initial screening process. Of these, 79 variants were identified as potentially carrying clinical relevance. Upon thorough validation by sanger sequencing of 111 variants, 26 (23.6%) of these loci were verified as wild-type sequences, while mutations were substantiated at 84 loci, among which 1 locus were recognized by 3 repeated probes. Due to discrepancies between the actual mutation statuses and the initial predictions by the ECMA platform, 34 SNVs were classified as either pathogenic (P) or likely pathogenic (LP), which constitutes 30.0% of all initially screened SNVs (Table S6). Considering genetic inheritance patterns and allelic compositions (homozygosity vs. heterozygosity), 17 SNVs were selected for their potential clinical significance (Table S7). Among the 12 fetuses with SNVs, two (Sample ID 323 and 324) were from monochorionic-diamniotic twin gestations.

Based on our results, two fetuses were identified with SNVs associated with deafness, and two were identified as having SNVs linked to Glucose-6-phosphate dehydrogenase (G6PD) deficiency. Additionally, pathologies associated with the remaining SNVs comprised Osteogenesis Imperfecta, Ornithine Transcarbamylase Deficiency, Hypercholesterolemia, Achondroplasia, and Mediterranean Anemia. Among the 13 fetuses with identified SNVs, follow-up was declined by the families of 4 fetuses, whilst 4 fetuses underwent termination of pregnancy based on the combination of SNV results, family clinical history, and ultrasound findings.

#### The detection yield of the comprehensive ECMA in fetal structural anomalies

These fetuses with structural anomalies were classified into 11 subgroups, including increased NT (28.8%, 49/170), multisystem (24.1%, 41/170), cardiac (12.4%, 21/170), renal (8.8%, 15/170), skeletal (7.6%, 13/170), brain (6.5%, 11/170), craniofacial (5.3%, 9/170), abdominal (2.9%, 5/170), fetal growth restriction (1.8%, 3/70), spinal (1.2%, 2/170), lymphatic or effusion (0.6%, 1/170).

The diagnostic rate differed among these phenotypic subgroups. NT (8.2%, 4/49), multisystem (34.1%, 14/41), cardiac (4.8%, 1/21), renal (6.7%, 1/15), skeletal (15.4%, 2/13), abdominal (20.0%, 1/5), fetal growth restriction, spinal (50.5%, 1/2). No genetic variants were detected in fetuses with lymphatic or effusion (n=1), brain (n=11), craniofacial anomalies(n=9). (see Figure 3)

**Figure 3.**
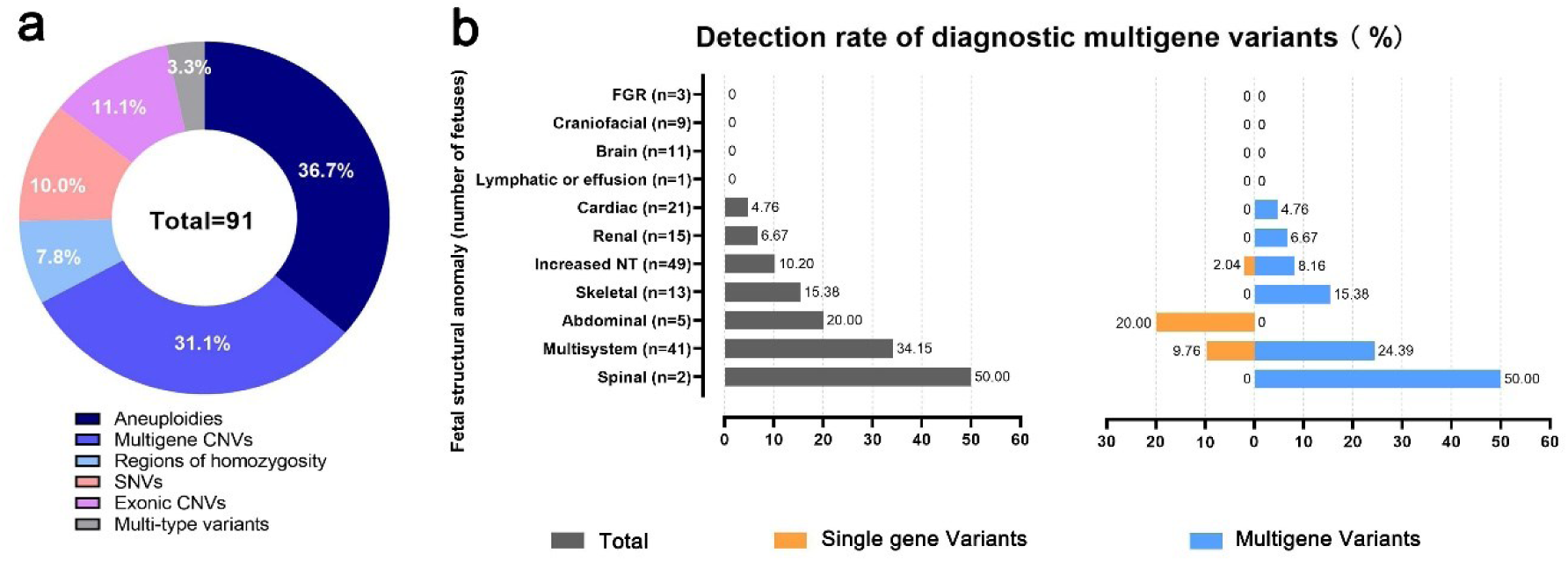
The detection rate of ECMA and diagnostic genetic variants in pregnancies complicated by fetal structural anomalies. (A)Genetic variants was identified in 91 (out of 512, 17.6%) pregnancies. Of these, 33 (36.7%) had aneuploidies, 28 had multigene copy number variations (CNV, 31.1%), 7 (7.8%) had regions of homozygosity, 9 (10.0%) had single nucleotide variants (SNVs) and 10 (11.1%) had exonic CNVs. Additionally, 3 cases exhibited multiple types of variant conditions. (B) In all 170 pregnancies complicated by fetal structural anomalies, the detection rate for genetic variants was assessed. Single gene variants comprise exonic CNVs and SNVs. Multigene variants encompass aneuploidies, multigene CNVs and regions of homozygosity.

## Discussion

### Principal Findings

The main finding of this study is that expanded chromosomal microarray identified genetic abnormalities in 91 out of 512 cases (17.6%). The encountered rate was significantly higher than the rates observed with low-depth GS (66 out of 512 cases, 12.9%) and conventional chromosome karyotyping (42 out of 512 cases, 8.2%). ECMA not only detected all these non-mosaic aneuploidies and copy number variations in 62(12.1%) diagnosed cases identified by low-depth GS, but also detected 9 cases with regions of homozygosity, 10(2.0%) cases with exonic deletions (*SMN1* and *DMD*), and 13(2.3%) cases with single nucleotide variations. The total diagnostic yield in our study was consistent with that reported in previous studies (19.8% and 18.9%, respectively).

### Results in the Context of What is Known

In comparison to the entire cohort of 512 individuals, the average maternal age of those carrying fetuses with aneuploidy was only marginally higher by 1.2 years, demonstrating a fundamental similarity. However, there was a notable increase in the proportion of mothers aged 35 years or older among those carrying aneuploid fetuses, with a prevalence of 51.6%, representing a 12% elevation compared to the overall dataset of 512 subjects.

The total diagnostic yield of ECMA in our study was consistent with that reported in previous studies using low-depth GS (12.2%) and CMA (11.8%)^10^. Compared to low-depth GS, CMA is capable of identifying ROH, yet it fails to detect cases of low-level mosaicism (<10%). The disparity in detection rates between these two techniques is contingent upon the number of low-level mosaicism and samples with ROH. In the context of prenatal applications, the detection rate of ECMA is consistent with that of whole genome sequencing (16.8%-19.8%)^36,37^.

In our study, the turnaround time (TAT) for ECMA was observed to be within 2 to 3 weeks. As ECMA is categorized under the recommended CMA protocols, it is offered to any patient opting for invasive diagnostic testing. Its application in the prenatal context showcases a commendable TAT, thereby augmenting clinical efficacy and facilitating patient care management.

### Clinical and Research Implications

The accurate detection of non-mosaic aneuploidies, such as Trisomy 21, Trisomy 18, Trisomy 13, Trisomy X, and Klinefelter syndrome, is of paramount importance for prenatal diagnosis and genetic counseling^38,39^.

In our study, CMA not only successfully detected all 31 CNVs identified via low-depth GS, but also detected additional exonic CNV (<100 Kb) that was beyond the detection range of low-depth GS. 8 individuals with deletion in the *SMN1* exon7 gene was identified. Whole Genome Sequencing (WGS) is considered the diagnostic approach with the highest detection rate currently; however, among the 642 reported cases of WGS application in prenatal clinical settings, no reporting has been observed specifically regarding SMA screening outcomes^36,37,40^.

Due to the patient cohort encompassing a wide variety of fetal abnormalities rather than targeting specific conditions, this study possesses enhanced generalizability, enabling it to reveal the detectability of ECMA screening for both chromosomal and monogenic variations. Although such screening cannot replace imaging-based screening procedures, it serves as a supplementary tool for the early identification of asymptomatic fetuses that may not be evident on routine prenatal ultrasound screening in the first trimester, such as those with achondroplasia. In the study, pathogenic variants associated with postnatal metabolic diseases and treatable conditions were identified in 9 out of 13 fetuses (69.2%) with monogenic diseases. The benefits of prenatal ECMA screening extend beyond the prenatal stage; postnatally, infants can greatly benefit from preventive measures such as tailored diets and regular check-ups^41–45^.

Among the 17 single nucleotide variants (SNVs) identified, the pathogenic variant c.1138G>A (p.Gly380Arg) in the *FGFR3* gene stands out as particularly significant. This specific mutation is found in approximately 98% of individuals with achondroplasia, highlighting its strong association with the condition^46^. Identifying a heterozygous pathogenic variant such as c.1138G>A in the *FGFR3* gene can effectively establish a diagnosis of achondroplasia.^47^ Moreover, prenatal testing is a viable option when a pregnancy presents an increased risk for achondroplasia due to the presence of such variants.^46^

The findings underscore the importance of integrating various diagnostic tools to achieve a thorough and accurate genetic assessment, which is crucial for effective genetic counseling and clinical management.

The ECMA methodology features an independent analysis platform and a standardized analysis protocol, thereby substantially reducing the personnel and analytical capacity requirements compared to WGS. Additionally, through synthesizing consensus from multiple stakeholders, this study has compiled a tailored screening gene list specific to the ECMA platform. Collectively, ECMA serve to mitigate the influence of individual operator variability on the analytical outcomes, thereby enhancing the reliability and consistency of the results obtained.

Prenatal diagnosis imposes stringent requirements on the TAT. Relative to traditional CMA, ECMA does not augment the TAT duration. In our study the TAT is 2-3 weeks, aligning with findings reported in existing literature^48^. Nonetheless, it entails a supplementary procedure for the validation of expanded results. Cost is another vital issue impacting the feasibility in clinical use. Aside from the supplementary validation steps necessitated by the expanded results, ECMA does not introduce incremented expenses when contrasted with the prevailing CMA methodology. Our cost (supplementary validation included) is US$ 500, aligning with findings reported in existing literature^48^.

### Strengths and Limitations

This study is a large prospective study focused on the application and utility analysis of ECMA which targets three of the most prevalent types of pathogenic genetic variants: aneuploidies, CNVs and monogenic variants in prenatal diagnosis. The ECMA assay identified 8 (out of 512) carriers of SMA and 13 (out of 512) SNVs potentially relevant to clinical presentations, all of which were undetectable by conventional CMA. Currently, no commercially available CNV detection method inclusive of SMA screening is in clinical application. While the assay demonstrates a satisfactory SNV detection rate, Sanger sequencing is required for validation, and it is noteworthy that the positive predictive value (PPV) for SNVs stands at only 20.5%.

## Conclusions

We provide further evidence that ECMA is a comprehensive diagnostic tool capable of detecting both non-mosaic and mosaic aneuploidies, with sensitivity extending to mosaicisms of over 20%, in addition to multigene CNVs, exonic CNVs, and facilitating screening for conditions such as SMA, DMD, and monogenic disorders. The study also underscores the clinical characteristics of the participants, thereby furnishing valuable demographic insights and elucidating the indications for prenatal and genetic testing within the studied cohort.

## Data Availability

All data produced in the present work are contained in the manuscript

## Acknowledgement

We thank all the patients who participated in this study

