## Supplemental figure and tables for "Expanded chromosomal microarray comprising screening for spinal muscular atrophy and monogenic diseases3"

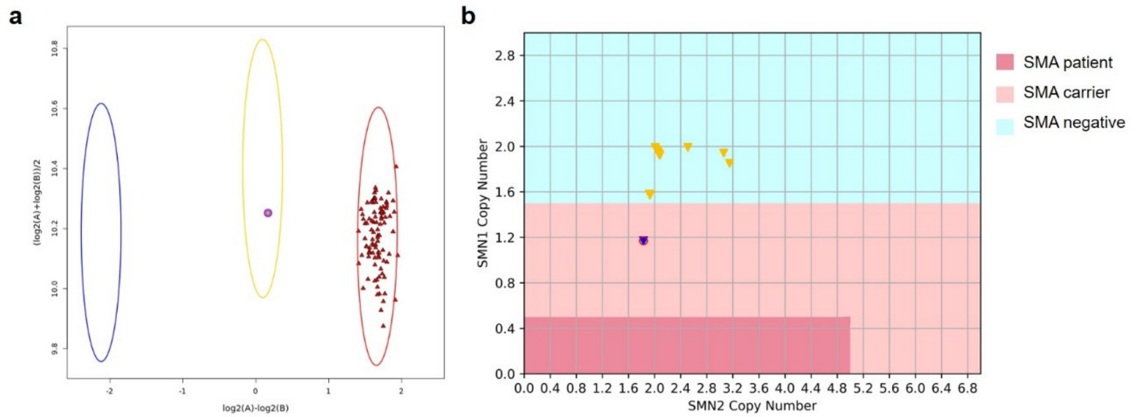


**Figure S1. Diagrams illustrating the interpretation of SNV and SMA test results.** (A) Interpretation of single nucleotide variants (SNVs) results. After cluster analysis, points within the blue circle are homozygotes, points within the yellow circle are heterozygote, points within the red circle are wild type SNVs. All analyzed samples of same array are shown as triangle. (B) Interpretation of *SMN1* and *SMN2* Copy Number Results. For Spinal Muscular Atrophy (SMA) patient with homozygous *SMN1* exon 7 deletion, results are 0 ≤ *SMN1* copy number ≤ 0.50, *SMN*2copy number< 5 (dark pink area). The outcomes for SMA carriers fall within the light pink region. For SMA carrier with homozygous *SMN1* exon 7 deletion, results are 0 ≤ *SMN1* copy number ≤0.50, *SMN*2 copy number ≥ 5. For SMA carrier with *SMN1* exon 7 heterozygous deletion, results are 0.5 ≤ *SMN1* copy number ≤ 1.50.

**Table S1. Results of ECMA, low-depth GS, and karyotype analysis in 512 fetuses**

| Clinical information | | | | low-depth GS results | Karyotype results | ECMA Results | | | |
|---|---|---|---|---|---|---|---|---|---|
| sample ID | Age (years) | Group * | Gestational age | | | Main Results | SMA screening results | *SMN1* CN | *SMN2* CN |
| 1 | 30-34 | 3, 7 | 20+2 | 46,XN | 46,XN | 46,XN | unaffected | 1.99 | 2.01 |
| 2 | 30-34 | 3, 7 | 17+3 | seq(X)×3 | 47,XXX | arr(X)×3 | unaffected | 1.85 | 1.15 |
| 3 | 25-29 | 7 | 17+6 | 46,XN | 46,XN | 46,XN | unaffected | 1.96 | 1.04 |
| 4 | 30-34 | 3, 6 | 17+4 | seq[hg19]dup(15)(q11.2q13.1) chr15:g.23620000_28420000dup | 46,XN | arr[hg19]15q11.2q13.1(23616115-28555716)×3 | unaffected | 1.94 | 1.06 |
| 5 | 30-34 | 6 | 25+2 | 46,XN | 46,XN | 46,XN | unaffected | 1.98 | 2.02 |
| 6 | 35-39 | 3, 4 | 18+2 | 46,XN | 46,XN | 46,XN | unaffected | 1.98 | 2.02 |
| 7 | 30-34 | 7, 8 | 16+6 | seq(21)×3[0.1] | 46,XN | 46,XN | unaffected | 1.96 | 1.04 |
| 8 | 30-34 | 3, 5, 8 | 18+3 | 46,XN | 46,XN | 46,XN | unaffected | 1.95 | 1.05 |
| 9 | 35-39 | 4, 6 | 23+2 | 46,XN | 46,XN | 46,XN | unaffected | 1.98 | 2.02 |
| 10 | 25-29 | 1, 3 | 17+2 | 46,XN | 46,XN | 46,XN | unaffected | 1.99 | 2.01 |
| 11 | 25-29 | 7 | 17+5 | 46,XN | 46,XN | 46,XN | unaffected | 2 | 2 |
| 12 | 30-34 | 3, 5, 8 | 17+1 | 46,XN | 46,XN | 46,XN | unaffected | 2.04 | 1.96 |
| 13 | 35-39 | 3, 4 | 19+2 | 46,XN | 46,XN,t(4;17)(q31.3;p11.2) | arr(11p)×2 hmz [0.5] | unaffected | 1.97 | 2.03 |
| 14 | 35-39 | 4 | 17+2 | 46,XN | 46,XN | 46,XN | unaffected | 1.93 | 1.07 |
| 15 | ≥40 | 4, 7 | 23+6 | seq(21)×3 | 47,XN,+21 | arr(21)×3 | unaffected | 2 | 2 |
| 16 | 30-34 | 3, 7 | 18+5 | 46,XN | 46,XN | 46,XN | unaffected | 1.97 | 2.03 |
| 17 | 30-34 | 6 | 27+4 | 46,XN | 46,XN | 46,XN | unaffected | 1.97 | 1.03 |
| 18 | 25-29 | 6 | 23+5 | 46,XN | 46,XN | 46,XN | unaffected | 1.97 | 2.03 |
| 19 | 25-29 | 6 | 24+1 | 46,XN | 46,XN | 46,XN | unaffected | 1.95 | 1.05 |
| 20 | 30-34 | 1, 3 | 17+5 | 46,XN | 46,XN | 46,XN | unaffected | 1.98 | 2.02 |
| 21 | ≤24 | 7 | 21+3 | 46,XN | 46,XN | 46,XN | unaffected | 1.99 | 2.01 |
| 22 | 35-39 | 3, 4 | 13+3 | 46,XN | 46,XN | 46,XN | unaffected | 1.94 | 2.06 |
| 23 | ≤24 | 6 | 12+5 | 46,XN | 46,XN | 46,XN | unaffected | 1.96 | 2.04 |
| 24 | 25-29 | 3, 5, 8 | 17+1 | 46,XN | 45，XN,der(13,14)(q10;q10)pat | 46,XN | unaffected | 1.92 | 2.08 |
| 25 | 25-29 | 7 | 24+4 | 46,XN | 46,XN | 46,XN | unaffected | 2.09 | 1.91 |
| 26 | 25-29 | 6 | 16+2 | 46,XN | 46,XN | 46,XN | unaffected | 1.92 | 1.08 |
| 27 | 25-29 | 1, 3, 7 | 19+2 | 46,XN | 46,XN | 46,XN | unaffected | 1.9 | 0.1 |
| 28 | 35-39 | 4, 7 | 19+5 | seq(21)×3[0.15] | 45,X[2]/46,XX[18] | 46,XN | unaffected | 1.95 | 1.05 |
| 29 | 25-29 | 6 | 16+3 | 46,XN | 46,XN | 46,XN | unaffected | 2 | 2 |
| 30 | 25-29 | 2, 3 | 18 | 46,XN | 46,XN | 46,XN | unaffected | 1.98 | 2.02 |
| 31 | 35-39 | 4 | 17+4 | 46,XN | 46,XN | 46,XN | unaffected | 1.94 | 1.06 |
| 32 | 30-34 | 3, 7 | 19+4 | 46,XN | 46,XN | 46,XN | unaffected | 2.03 | 1.97 |
| 33 | ≥40 | 3, 4, 8 | 17+5 | 46,XN | 46,XN | 46,XN | unaffected | 1.92 | 1.08 |
| 34 | 30-34 | 3 | 17+6 | 46,XN | 46,XN | 46,XN | unaffected | 1.93 | 1.07 |
| 35 | 25-29 | 2, 3, 7 | 21+3 | 46,XN | 46,XN | 46,XN | unaffected | 1.86 | 0.14 |
| 36 | 25-29 | 6 | 30+5 | 46,XN | 46,XN | 46,XN | unaffected | 1.96 | 1.04 |
| 37 | ≥40 | 3, 4, 8 | 16+5 | 46,XN | 46,XN | 46,XN | unaffected | 2.29 | 2.21 |
| 38 | 30-34 | 3, 7 | 19+4 | 46,XN | 46,XN | 46,XN | unaffected | 2.02 | 1.98 |
| 39 | 30-34 | 3, 7 | 19+4 | 46,XN | 46,XN | 46,XN | unaffected | 1.98 | 2.02 |
| 40 | 25-29 | 6 | 18+2 | 46,XN | 46,XN | 46,XN | unaffected | 2.86 | 1.14 |
| 41 | 25-29 | 7 | 20+1 | 46,XN | 46,XN | 46,XN | unaffected | 1.99 | 2.01 |
| 42 | 25-29 | 7 | 22+3 | 46,XN | 46,XN | 46,XN | unaffected | 1.94 | 1.06 |
| 43 | 35-39 | 4 | 18+1 | 46,XN | 46,XN | 46,XN | unaffected | 1.97 | 1.03 |
| 44 | 35-39 | 4 | 24+5 | 46,XN | 46,XN | 46,XN | unaffected | 1.95 | 1.05 |
| 45 | 35-39 | 3, 4 | 16+2 | 46,XN | 46,XN | 46,XN | unaffected | 1.93 | 1.07 |
| 46 | 30-34 | 6 | 29+5 | 46,XN | 46,XN | 46,XN | unaffected | 2.04 | 1.96 |
| 47 | 35-39 | 3, 4, 6 | 16 | seq(21)×3 | 47,XN,+21 | arr(21)×3 | unaffected | 1.98 | 2.02 |
| 48 | 35-39 | 3, 4 | 17+1 | 46,XN | 46,XN | 46,XN | unaffected | 2.85 | 2.15 |
| 49 | 30-34 | 6 | 16+3 | 46,XN | 46,XN | 46,XN | unaffected | 2.02 | 1.98 |
| 50 | ≥40 | 3, 4 | 17 | 46,XN | 46,XN | 46,XN | unaffected | 2 | 2 |
| 51 | 30-34 | 3, 6, 7 | 22+6 | 46,XN | 46,XN | 46,XN | unaffected | 2.01 | 1.99 |
| 52 | 35-39 | 4, 8 | 17+5 | 46,XN | 46,XN | 46,XN | unaffected | 1.91 | 0.09 |
| 53 | 35-39 | 4, 6 | 22+6 | 46,XN | 46,XN | 46,XN | unaffected | 1.98 | 2.02 |
| 54 | 30-34 | 3, 7 | 18+5 | seq(21)×3 | 47,XN,+21 | arr(21)×3 | unaffected | 1.9 | 1.1 |
| 55 | 25-29 | 7 | 17+6 | 46,XN | 46,XN | 46,XN | unaffected | 1.98 | 2.02 |
| 56 | 30-34 | 3, 6 | 24+6 | 46,XN | 46,XN | 46,XN | unaffected | 1.63 | 1.87 |
| 57 | 35-39 | 3, 4, 6 | 22 | 46,XN | 46,XN | 46,XN | unaffected | 1.9 | 2.1 |
| 58 | 30-34 | 3, 7 | 18 | 46,XN | 46,XN | 46,XN | unaffected | 2.04 | 1.96 |
| 59 | 35-39 | 3, 4 | 26+3 | seq[hg19]dup(1)(q44) chr1:g.246220000_247500000dup | 46,XN | arr[hg19]1q44(246217629-247516609)×3 | unaffected | 1.98 | 1.02 |
| 60 | 30-34 | 7 | 16+1 | seq(X)×1[0.2] | 45,X[4]/46,XX[26] | 46,XN | unaffected | 1.89 | 2.11 |
| 61 | ≥40 | 4 | 18 | 46,XN | 46,XN | 46,XN | unaffected | 2 | 2 |
| 62 | ≥40 | 3, 4 | 18+2 | 46,XN | 46,XN | 46,XN | unaffected | 1.98 | 2.02 |
| 63 | 35-39 | 4 | 18+2 | seq[hg19]del(4)(q22.1q22.3) chr4:g.93300000_96960000del | 46,XN | arr[hg19]4q22.1q22.3(93296704-97008926)×1 | unaffected | 1.99 | 1.01 |
| 64 | ≤24 | 6 | 26+1 | 46,XN | 46,XN | 46,XN | unaffected | 1.97 | 2.03 |
| 65 | 35-39 | 1, 4 | 15 | 46,XN | 46,XN | 46,XN | unaffected | 1.99 | 2.01 |
| 66 | 30-34 | 3, 6 | 21+6 | 46,XN | 46,XN | 46,XN | unaffected | 1.93 | 2.07 |
| 67 | 35-39 | 3, 4 | 17+5 | 46,XN | 46,XN | 46,XN | unaffected | 1.98 | 2.02 |
| 68 | 35-39 | 3, 4, 6 | 25 | 46,XN | 46,XN | 46,XN | unaffected | 1.94 | 1.06 |
| 69 | 25-29 | 3, 6 | 23+3 | 46,XN | 46,XN | 46,XN | unaffected | 1.94 | 1.06 |
| 70 | ≥40 | 4 | 16+1 | 46,XN | 46,XN | 46,XN | unaffected | 2 | 2 |
| 71 | 35-39 | 3, 4 | 18+1 | 46,XN | 46,XN | 46,XN | unaffected | 1.92 | 1.08 |

| Clinical information | | | | low-depth GS results | Karyotype results | ECMA Results | | | |
|---|---|---|---|---|---|---|---|---|---|
| sample ID | Age (years) | Group * | Gestational age | | | Main Results | SMA screening results | *SMN1* CN | *SMN2* CN |
| 72 | 30-34 | 7 | 19+5 | 46,XN | 46,XN | 46,XN | unaffected | 1.99 | 2.01 |
| 73 | 30-34 | 7 | 21+1 | 46,XN | 46,XN | 46,XN | unaffected | 1.91 | 1.09 |
| 74 | 35-39 | 4, 6 | 23+1 | 46,XN | 46,XN | 46,XN | unaffected | 1.87 | 2.13 |
| 75 | 25-29 | 3, 7 | 17+4 | 46,XN | 46,XN | 46,XN | unaffected | 1.98 | 2.02 |
| 76 | 30-34 | 6 | 26+3 | 46,XN | 46,XN | 46,XN | unaffected | 1.64 | 1.36 |
| 77 | 35-39 | 3, 4 | 19+4 | 46,XN | 46,XN | 46,XN | unaffected | 2.01 | 1.99 |
| 78 | ≤24 | 6 | 13+1 | seq(X)×1 | 45,X | arr(X)×1 | unaffected | 1.95 | 1.05 |
| 79 | 25-29 | 1, 2, 3 | 19+3 | 46,XN | 46,XN | 46,XN | unaffected | 2.02 | 1.98 |
| 80 | 35-39 | 3, 4 | 13+3 | 46,XN | 46,XN | 46,XN | unaffected | 2 | 2 |
| 81 | 30-34 | 3 | 18+2 | 46,XN | 46,XN | 46,XN | unaffected | 2 | 2 |
| 82 | 35-39 | 3, 4, 5, 8 | 16+5 | seq[hg19]dup(11)(q21q22.1) chr11:g.95740000_98300000dup | 46,XN,t(4;8)(q35;q13) mat | arr[hg19]11q21q22.1(95726001-98289956)×3 | unaffected | 1.98 | 2.02 |
| 83 | ≥40 | 2, 3, 4 | 17 | 46,XN | 46,XN | 46,XN | unaffected | 1.93 | 2.07 |
| 84 | 30-34 | 9 | 17 | 46,XN | 46,XN,15pstk+ | 46,XN | unaffected | 2 | 2 |
| 85 | 35-39 | 4 | 22+6 | 46,XN | 46,XN | 46,XN | unaffected | 1.91 | 1.09 |
| 86 | 30-34 | 6 | 22+6 | 46,XN | 46,XN | 46,XN | unaffected | 1.94 | 1.06 |
| 87 | 25-29 | 6 | 19+3 | 46,XN | 46,XN,16qh+ | 46,XN | unaffected | 1.99 | 2.01 |
| 88 | 25-29 | 6 | 19+3 | 46,XN | 46,XN,16qh+ | 46,XN | unaffected | 1.96 | 2.04 |
| 89 | ≥40 | 4 | 18+2 | 46,XN | 46,XN | 46,XN | unaffected | 2.22 | 2.28 |
| 90 | ≥40 | 4 | 18+2 | 46,XN | 46,XN | 46,XN | unaffected | 1.92 | 1.08 |
| 91 | 30-34 | 9 | 22 | 46,XN | 46,XN | 46,XN | unaffected | 1.98 | 2.02 |
| 92 | 25-29 | 3, 6 | 21+6 | 46,XN | 46,XN | 46,XN | unaffected | 1.98 | 2.02 |
| 93 | ≥40 | 4, 5, 8 | 16+1 | 46,XN | 46,XN | 46,XN | unaffected | 1.97 | 1.03 |
| 94 | 35-39 | 3, 4 | 17+5 | 46,XN | 46,XN | 46,XN | unaffected | 1.95 | 2.05 |
| 95 | 30-34 | 5 | 18+1 | 46,XN | 46,XN | 46,XN | unaffected | 1.93 | 1.07 |
| 96 | 30-34 | 6 | 24+2 | 46,XN | 46,XN | 46,XN | unaffected | 1.98 | 2.02 |
| 97 | 30-34 | 5 | 17 | 46,XN | 46,XN | 46,XN | unaffected | 2 | 2 |
| 98 | ≥40 | 4 | 17+3 | 46,XN | 46,XN | 46,XN | unaffected | 1.93 | 2.07 |
| 99 | ≤24 | 9 | 21+5 | 46,XN | 46,XN | 46,XN | unaffected | 1.98 | 2.02 |
| 100 | 25-29 | 3, 6 | 22+2 | 46,XN | 46,XN | 46,XN | unaffected | 2.18 | 1.82 |
| 101 | 30-34 | 6 | 28 | 46,XN | 46,XN | 46,XN | unaffected | 1.99 | 2.01 |
| 102 | 25-29 | 6 | 22+4 | 46,XN | 46,XN | 46,XN | unaffected | 1.94 | 1.06 |
| 103 | 30-34 | 7 | 19+4 | 46,XN | 46,XN | 46,XN | unaffected | 1.93 | 1.07 |
| 104 | 35-39 | 3, 4, 6 | 22+4 | 46,XN | 46,XN | 46,XN | unaffected | 1.95 | 1.05 |
| 105 | 30-34 | 3, 6 | 24+4 | 46,XN | 46,XN | 46,XN | unaffected | 1.92 | 1.08 |
| 106 | 30-34 | 3, 5, 7 | 21+1 | 46,XN | 46,XN | 46,XN | unaffected | 1.93 | 1.07 |
| 107 | ≥40 | 4 | 17+3 | 46,XN | 46,XN | 46,XN | unaffected | 1.96 | 2.04 |
| 108 | 30-34 | 3, 5, 8 | 18+4 | 46,XN | 46,XN， t(2;4)(p22;q27) mat | 46,XN | unaffected | 1.98 | 1.02 |
| 109 | 25-29 | 2, 3 | 17+1 | 46,XN | 46,XN | 46,XN | unaffected | 1.95 | 1.05 |
| 110 | 30-34 | 1, 5 | 17+6 | 46,XN | 46,XN | 46,XN | unaffected | 1.99 | 1.01 |
| 111 | 25-29 | 7 | 18+6 | seq(21)×3 | 47,XN,+21 | arr(21)×3 | unaffected | 1.92 | 2.08 |
| 112 | 30-34 | 3, 6 | 27+6 | 46,XN | 46,XN | 46,XN | unaffected | 1.98 | 2.02 |
| 113 | 30-34 | 3, 7 | 19+3 | 46,XN | 46,XN | 46,XN | unaffected | 2.02 | 1.98 |
| 114 | ≥40 | 4 | 17+5 | 46,XN | 46,XN | 46,XN | unaffected | 1.96 | 1.04 |
| 115 | 25-29 | 3, 6 | 17 | 46,XN | 46,XN | 46,XN | unaffected | 1.98 | 2.02 |
| 116 | ≥40 | 3, 4 | 22 | 46,XN | 46,XN | 46,XN | unaffected | 1.9 | 1.1 |
| 117 | 30-34 | 7 | 19+1 | 46,XN | 46,XN | 46,XN | unaffected | 1.97 | 1.03 |
| 118 | 25-29 | 6 | 23+6 | 46,XN | 46,XN | 46,XN | unaffected | 1.95 | 1.05 |
| 119 | 30-34 | 3, 7 | 18 | 46,XN | 46,XN | 46,XN | unaffected | 1.98 | 2.02 |
| 120 | ≤24 | 6 | 22+5 | 46,XN | 46,XN | 46,XN | unaffected | 1.99 | 2.01 |
| 121 | 35-39 | 4 | 16+2 | 46,XN | 46,XN | 46,XN | unaffected | 2.01 | 1.99 |
| 122 | 30-34 | 7 | 18+3 | 46,XN | 46,XN | 46,XN | unaffected | 1.93 | 2.57 |
| 123 | 35-39 | 4 | 17+1 | 46,XN | 46,XN | 46,XN | unaffected | 1.98 | 2.02 |
| 124 | 25-29 | 3, 6 | 22+6 | 46,XN | 46,XN | 46,XN | unaffected | 2 | 2 |
| 125 | 25-29 | 6 | 19+4 | 46,XN | 46,XN | 46,XN | unaffected | 1.91 | 1.09 |
| 126 | 30-34 | 7 | 18+2 | 46,XN | 46,XN | 46,XN | unaffected | 1.96 | 2.04 |
| 127 | 30-34 | 3 | 16+2 | 46,XN | 45,XN,rob(13;14)(q10; q10) | 46,XN | unaffected | 1.93 | 2.07 |
| 128 | 35-39 | 3, 4 | 17+3 | 46,XN | 46,XN | 46,XN | unaffected | 1.94 | 1.06 |
| 129 | 30-34 | 3, 6 | 24+4 | 46,XN | 46,XN | 46,XN | unaffected | 1.87 | 0.13 |
| 130 | ≥40 | 4 | 16+5 | 46,XN | 46,XN | 46,XN | unaffected | 2.01 | 1.99 |
| 131 | 30-34 | 7 | 18+3 | 46,XN | 46,XN | 46,XN | unaffected | 1.97 | 2.03 |
| 132 | ≥40 | 3, 4 | 18+3 | 46,XN | 46,XN | 46,XN | unaffected | 1.96 | 2.54 |
| 133 | 30-34 | 3, 8 | 18+1 | 46,XN | 46,XN | 46,XN | unaffected | 1.98 | 2.02 |
| 134 | 25-29 | 7 | 17+2 | seq(X)×3 | 47,XXX | arr(X)×3 | unaffected | 2.02 | 1.98 |
| 135 | 25-29 | 3, 7 | 21+5 | seq[hg19]del(X)(p22.12p22.11) chrX:g.20500000_23860000del | 46,XN | arr[hg19]Xp22.12p22.11(20536377-23832515)×1 | unaffected | 2.98 | 1.02 |
| 136 | 25-29 | 3, 8 | 17+6 | 46,XN | 46,XN | 46,XN | unaffected | 1.95 | 1.05 |
| 137 | 30-34 | 2, 3 | 17+1 | 46,XN | 46,XN | 46,XN | unaffected | 1.98 | 2.02 |
| 138 | 25-29 | 7 | 25+6 | seq[hg19]dup(13)(q21.31q34) chr13:g.64440000_115100000dup | 47,XN,+der(13)del(13)(q11q21.3) | arr[hg19]13q21.31q34(64407224-114800246)×3 | unaffected | 1.61 | 1.89 |
| 139 | 35-39 | 4 | 16+4 | 46,XN | 46,XN | 46,XN | unaffected | 1.9 | 0.1 |
| 140 | 30-34 | 3, 6 | 24+1 | 46,XN | 46,XN | 46,XN | unaffected | 2 | 1 |
| 141 | 30-34 | 3, 5, 8 | 18+6 | 46,XN | 46,XN,t(9;20)(p13;p11.2)pat | 46,XN | unaffected | 1.93 | 1.07 |
| 142 | 35-39 | 3, 4 | 16+3 | 46,XN | 46,XN | 46,XN | unaffected | 1.89 | 0.11 |
| 143 | 35-39 | 4, 7 | 19+4 | 46,XN | 46,XN | 46,XN | unaffected | 1.94 | 1.56 |

| Clinical information | | | | low-depth GS results | Karyotype results | ECMA Results | | | |
|---|---|---|---|---|---|---|---|---|---|
| sample ID | Age (years) | Group * | Gestational age | | | Main Results | SMA screening results | *SMN1* CN | *SMN2* CN |
| 144 | 35-39 | 4, 6 | 29+1 | 46,XN | 46,XN | 46,XN | unaffected | 1.98 | 2.02 |
| 145 | 35-39 | 4 | 17 | 46,XN | 46,XN | 46,XN | unaffected | 2 | 2 |
| 146 | 25-29 | 6 | 23+2 | 46,XN | 46,XN | 46,XN | unaffected | 2.08 | 0.92 |
| 147 | 30-34 | 5, 8 | 17+6 | 46,XN | 45,XN,rob(13;14)(q10;q10)pat | 46,XN | unaffected | 1.98 | 2.02 |
| 148 | 30-34 | 6 | 22+4 | 46,XN | 46,XN | 46,XN | unaffected | 1.88 | 1.12 |
| 149 | 30-34 | 3, 6 | 22+2 | 46,XN | 46,XN | 46,XN | unaffected | 1.95 | 1.05 |
| 150 | 30-34 | 3, 7 | 18+1 | 46,XN | 46,XN | 46,XN | unaffected | 1.96 | 2.04 |
| 151 | 30-34 | 6 | 23+2 | 46,XN | 46,XN | 46,XN | unaffected | 1.98 | 2.02 |
| 152 | 35-39 | 3, 4 | 17+3 | 46,XN | 46,XN | 46,XN | carrier | 1.18 | 2.82 |
| 153 | 25-29 | 3, 6 | 21+3 | 46,XN | 46,XN | 46,XN | unaffected | 1.96 | 2.04 |
| 154 | 25-29 | 3, 7 | 18+5 | 46,XN | 46,XN | 46,XN | unaffected | 2.02 | 1.98 |
| 155 | 35-39 | 3, 4 | 17+4 | 46,XN | 46,XN | 46,XN | unaffected | 2.05 | 0.95 |
| 156 | 30-34 | 6 | 22+1 | 46,XN | 46,XN | 46,XN | unaffected | 1.98 | 1.02 |
| 157 | 30-34 | 5, 8 | 17+5 | 46,XN | 46,XN,t(1;8)(q21;q23)pat | 46,XN | unaffected | 1.93 | 1.07 |
| 158 | 35-39 | 4, 6 | 23+2 | 46,XN | 46,XN | 46,XN | unaffected | 2.01 | 1.99 |
| 159 | 30-34 | 7 | 19+3 | 46,XN | 46,XN | 46,XN | unaffected | 2.04 | 1.96 |
| 160 | 35-39 | 4, 5, 8 | 21+3 | 46,XN | 46,XN | 46,XN | unaffected | 1.99 | 2.01 |
| 161 | ≥40 | 3, 4 | 19 | 46,XN | 46,XN | 46,XN | unaffected | 1.94 | 1.06 |
| 162 | 35-39 | 3, 4, 8 | 18+1 | 46,XN | 46,XN | 46,XN | unaffected | 3.06 | 0.94 |
| 163 | 35-39 | 4 | 18 | 46,XN | 46,XN | 46,XN | unaffected | 1.96 | 2.04 |
| 164 | 35-39 | 1, 3, 4 | 19 | 46,XN | 46,XN | 46,XN | unaffected | 1.87 | 1.13 |
| 165 | 30-34 | 3, 7 | 19+3 | 46,XN | 46,XN | 46,XN | unaffected | 1.96 | 2.04 |
| 166 | 35-39 | 3, 4 | 17+1 | 46,XN | 46,XN | 46,XN | unaffected | 2 | 2 |
| 167 | 35-39 | 3, 4, 6 | 17+2 | 46,XN | 46,X,inv(Y)(p11.2q11.23) | 46,XN | unaffected | 1.61 | 1.89 |
| 168 | ≥40 | 4 | 19 | 46,XN | 46,XN | 46,XN | unaffected | 1.99 | 2.01 |
| 169 | ≤24 | 7 | 17+3 | 46,XN | 46,XN | 46,XN | unaffected | 3.05 | 0.95 |
| 170 | 30-34 | 6 | 21 | 46,XN | 46,XN | 46,XN | unaffected | 2 | 2 |
| 171 | 30-34 | 2, 3 | 16+5 | 46,XN | 46,XN | 46,XN | unaffected | 1.92 | 1.08 |
| 172 | 25-29 | 7 | 19+1 | 46,XN | 46,XN | 46,XN | unaffected | 2.02 | 1.98 |
| 173 | 25-29 | 7 | 19+2 | 46,XN | 46,XN | 46,XN | unaffected | 2.03 | 1.97 |
| 174 | 35-39 | 4 | 17 | 46,XN | 46,XN | 46,XN | unaffected | 1.91 | 1.09 |
| 175 | 25-29 | 3, 6 | 23+2 | 46,XN | 46,XN | 46,XN | unaffected | 1.99 | 2.01 |
| 176 | 30-34 | 7 | 16+6 | 46,XN | 46,XN | 46,XN | unaffected | 2.13 | 0.87 |
| 177 | 30-34 | 5 | 17+2 | 46,XN | 46,XN | 46,XN | unaffected | 1.92 | 1.08 |
| 178 | 30-34 | 3, 7 | 19 | 46,XN | 46,XN | 46,XN | unaffected | 1.9 | 1.1 |
| 179 | 25-29 | 3, 6 | 21+3 | seq[hg19]del(12)(p13.33p13.31) chr12:g.160000_5880000del seq[hg19]dup(15)(q22.31q26.3) chr15:g.63720000_102400000dup | 46，XN,add(12)(p13)[25]/46,XN[5] | arr[hg19]12p13.33p13.31(83711-5897339)×1 arr[hg19]15q22.32q26.1(67202258-89337616)×3 | unaffected | 1.96 | 2.04 |
| 180 | 25-29 | 3, 7 | 17+3 | 46,XN | 46,XN | 46,XN | unaffected | 1.63 | 1.87 |
| 181 | 30-34 | 3, 5 | 18 | 46,XN | 46,XN | 46,XN | unaffected | 1.64 | 1.86 |
| 182 | 25-29 | 6 | 23+6 | 46,XN | 46,XN | 46,XN | unaffected | 1.96 | 1.04 |
| 183 | 25-29 | 7 | 18+6 | 46,XN | 46,XN | 46,XN | unaffected | 2.11 | 1.89 |
| 184 | ≤24 | 7 | 20+1 | seq[hg19]dup(22)(q11.21q11.23) chr22:g.18880000_25180000dup | 46,XN | arr[hg19]22q11.21q11.23(18614445-25127865)×3 | unaffected | 1.94 | 1.06 |
| 185 | 25-29 | 1 | 18+6 | 46,XN | 46,XN | 46,XN | unaffected | 1.92 | 1.08 |
| 186 | ≤24 | 7 | 19+1 | 46,XN | 46,XN | 46,XN | unaffected | 1.99 | 2.01 |
| 187 | ≤24 | 6 | 24 | 46,XN | 46,XN | 46,XN | unaffected | 1.6 | 0.4 |
| 188 | 35-39 | 4 | 16+6 | seq[hg19]del(15)(q13.3) chr15:g.32020000_32520000del | 46,XN | arr[hg19]15q13.3(32014503-32444185)×1 | unaffected | 3 | 1 |
| 189 | ≥40 | 3, 4, 8 | 18+3 | 46,XN | 46,XN | 46,XN | unaffected | 1.98 | 2.02 |
| 190 | 25-29 | 6 | 24+6 | 46,XN | 46,XN | 46,XN | unaffected | 1.96 | 2.04 |
| 191 | 25-29 | 7 | 20+4 | 46,XN | 46,XN | 46,XN | unaffected | 3.03 | 0.97 |
| 192 | 30-34 | 6 | 24+5 | 46,XN | 46,XN | 46,XN | unaffected | 2.02 | 1.98 |
| 193 | 35-39 | 3, 4 | 17+1 | 46,XN | 46,XN | 46,XN | carrier | 1.18 | 1.82 |
| 194 | ≥40 | 4 | 16+4 | 46,XN | 46,XN | 46,XN | unaffected | 2.01 | 1.99 |
| 195 | 35-39 | 3, 4 | 18+3 | 46,XN | 46,XN | 46,XN | unaffected | 2.01 | 1.99 |
| 196 | 25-29 | 6 | 23+4 | 46,XN | 46,XN | 46,XN | unaffected | 1.94 | 2.06 |
| 197 | 35-39 | 3, 4 | 17+4 | 46,XN | 46,XN | 46,XN | unaffected | 2.03 | 1.97 |
| 198 | 35-39 | 3, 4 | 18+4 | 46,XN | 46,XN | 46,XN | unaffected | 1.95 | 3.05 |
| 199 | ≤24 | 7 | 23+5 | 46,XN | 46,XN | 46,XN | unaffected | 1.98 | 2.02 |
| 200 | ≥40 | 4, 6 | 20+1 | 46,XN | 46,XN | 46,XN | unaffected | 3.11 | 0.89 |
| 201 | 30-34 | 6 | 13+2 | seq(21)×3 | 47,XN,+21 | arr(21)×3 | unaffected | 2 | 2 |
| 202 | 35-39 | 4, 6 | 23 | 46,XN | 46,XN | 46,XN | unaffected | 1.92 | 0.08 |
| 203 | 25-29 | 6 | 17+6 | seq(21)×3 | 47,XN,+21 | arr(21)×3 | unaffected | 2.02 | 1.98 |
| 204 | 25-29 | 7 | 18+6 | 46,XN | 46,XN | 46,XN | unaffected | 1.95 | 2.05 |
| 205 | ≥40 | 3, 4 | 18 | 46,XN | 46,XN | 46,XN | unaffected | 1.96 | 2.04 |
| 206 | 30-34 | 3, 6 | 24+1 | 46,XN | 46,XN | 46,XN | unaffected | 2.46 | 2.54 |
| 207 | 25-29 | 3, 7 | 22+1 | seq[hg19]del(X)(p22.31) chrX:g.6440000_8140000del | 46,XN | arr[hg19]Xp22.31(6454813-8126718)×0 | unaffected | 1.95 | 1.05 |
| 208 | ≥40 | 4 | 17+5 | seq(21)×3 | 47,XN,+21 | arr(21)×3 | unaffected | 2 | 2 |
| 209 | 30-34 | 6 | 23+3 | 46,XN | 46,XN | 46,XN | unaffected | 1.96 | 2.04 |
| 210 | 30-34 | 3, 5 | 16+4 | 46,XN | 46,XN | 46,XN | unaffected | 1.96 | 2.04 |
| 211 | 25-29 | 6 | 16+6 | 46,XN | 46,XN | 46,XN | unaffected | 1.61 | 1.89 |
| 212 | 35-39 | 4, 6 | 23+4 | 46,XN | 46,XN | 46,XN | unaffected | 1.99 | 2.01 |

| Clinical information | | | | low-depth GS results | Karyotype results | ECMA Results | | | |
|---|---|---|---|---|---|---|---|---|---|
| sample ID | Age (years) | Group * | Gestational age | | | Main Results | SMA screening results | *SMN1* CN | *SMN2* CN |
| 213 | 25-29 | 6 | 24+2 | 46,XN | 46,XN | 46,XN | carrier | 1.02 | 1.98 |
| 214 | 35-39 | 4 | 18+2 | 46,XN | 46,XN | 46,XN | unaffected | 2 | 2 |
| 215 | 25-29 | 3, 7 | 20+5 | 46,XN | 46,XN | 46,XN | unaffected | 1.99 | 2.01 |
| 216 | 35-39 | 3, 4 | 17+5 | 46,XN | 46,XN | 46,XN | unaffected | 1.96 | 2.04 |
| 217 | 30-34 | 9 | 25+3 | 46,XN | 46,XN | 46,XN | unaffected | 2.04 | 1.96 |
| 218 | 25-29 | 6, 7 | 28+3 | 46,XN | 46,XN | 46,XN | unaffected | 1.91 | 0.09 |
| 219 | ≥40 | 4, 6, 7 | 16+5 | 46,XN | 46,XN | 46,XN | unaffected | 1.99 | 2.01 |
| 220 | 30-34 | 3, 6 | 23+6 | 46,XN | 46,XN | 46,XN | unaffected | 1.9 | 0.1 |
| 221 | ≤24 | 3, 6 | 23 | 46,XN | 46,XN | 46,XN | unaffected | 1.95 | 2.05 |
| 222 | 35-39 | 4, 6 | 16+2 | 46,XN | 46,XN | 46,XN | unaffected | 1.7 | 2.8 |
| 223 | 35-39 | 4, 7 | 28 | seq(21)×3 | 47,XN,+21 | arr(21)×3 | carrier | 1.16 | 1.84 |
| 224 | 25-29 | 9 | 18+4 | 46,XN | 46,XN | 46,XN | unaffected | 1.99 | 2.01 |
| 225 | 35-39 | 3, 4, 7 | 16+3 | seq(21)×3 | 47,XN,+21 | arr(21)×3 | unaffected | 1.87 | 2.13 |
| 226 | ≥40 | 3, 4 | 17+1 | 46,XN | 46,XN | 46,XN | unaffected | 1.94 | 1.06 |
| 227 | 30-34 | 3, 6 | 17+5 | 46,XN | 46,XN | 46,XN | unaffected | 1.97 | 2.03 |
| 228 | 30-34 | 7 | 20+2 | seq[hg19]del(X)(q24) chrX:g.118960000_119100000del | 46,XN | arr[hg19]Xq24(118940712-119104408)×1 | unaffected | 1.98 | 1.02 |
| 229 | 30-34 | 7 | 18+3 | 46,XN | 46,XN | 46,XN | unaffected | 1.92 | 1.08 |
| 230 | 35-39 | 3, 4, 6 | 18+5 | 46,XN | 46,XN | 46,XN | unaffected | 1.97 | 2.03 |
| 231 | 35-39 | 3, 4, 6 | 18+5 | 46,XN | 46,XN | 46,XN | unaffected | 1.9 | 2.1 |
| 232 | 30-34 | 6 | 23+3 | 46,XN | 46,XN | 46,XN | unaffected | 1.9 | 2.1 |
| 233 | 30-34 | 6 | 23+3 | 46,XN | 46,XN | 46,XN | unaffected | 1.97 | 2.03 |
| 234 | 25-29 | 6 | 17+6 | 46,XN | 46,XN | 46,XN | unaffected | 1.99 | 2.01 |
| 235 | 25-29 | 6 | 17+6 | 46,XN | 46,XN | 46,XN | unaffected | 2.03 | 1.97 |
| 236 | 35-39 | 3, 4 | 17+2 | 46,XN | 46,XN | 46,XN | unaffected | 1.95 | 1.05 |
| 237 | 25-29 | 3, 6 | 22+5 | 46,XN | 46,XN | 46,XN | unaffected | 1.97 | 1.03 |
| 238 | 35-39 | 1, 4 | 17+1 | 46,XN | 46,XN | 46,XN | unaffected | 1.92 | 1.08 |
| 239 | 35-39 | 4 | 16+5 | 46,XN | 46,XN | 46,XN | unaffected | 1.91 | 1.09 |
| 240 | 35-39 | 2, 3, 4 | 17+4 | 46,XN | 46,XN | 46,XN | unaffected | 2.02 | 0.98 |
| 241 | 30-34 | 3, 6 | 25+4 | 46,XN | 46,XN | 46,XN | unaffected | 2.08 | 1.92 |
| 242 | 35-39 | 3, 4 | 17 | 46,XN | 46,XN | 46,XN | carrier | 1.06 | 0.94 |
| 243 | 35-39 | 3, 4 | 16+6 | 46,XN | 46,XN | 46,XN | unaffected | 1.98 | 2.02 |
| 244 | 35-39 | 3, 4, 7 | 19+6 | seq(X)×2,(Y)×1 | 47,XXY | arr(X)×2,(Y)×1 | unaffected | 1.95 | 2.05 |
| 245 | 35-39 | 4 | 16+4 | 46,XN | 46,XN | 46,XN | unaffected | 1.98 | 2.02 |
| 246 | 35-39 | 3, 4 | 17+3 | 46,XN | 46,XN | 46,XN | unaffected | 1.97 | 2.03 |
| 247 | ≥40 | 4 | 16+3 | 46,XN | 46,XN | 46,XN | unaffected | 2 | 2 |
| 248 | 25-29 | 6 | 23 | 46,XN | 46,XN | 46,XN | unaffected | 2.03 | 1.97 |
| 249 | 30-34 | 9 | 24+4 | 46,XN | 46,XN,t(5;19)(q11.1;q11.1) | 46,XN | unaffected | 2 | 2 |
| 250 | 35-39 | 4, 6 | 23+6 | seq[hg19]del(11)(p14.3) chr11:g.23320000_24680000del | 46,XN | arr[hg19]11p14.3(23337011-24681532)×1 | unaffected | 1.95 | 0.05 |
| 251 | 25-29 | 3, 6 | 24+2 | 46,XN | 46,XN | 46,XN | unaffected | 1.98 | 2.02 |
| 252 | 25-29 | 6 | 22 | 46,XN | 46,XN | 46,XN | unaffected | 2.05 | 1.95 |
| 253 | 35-39 | 4 | 18+4 | 46,XN | 46,XN | 46,XN | unaffected | 2 | 2 |
| 254 | 30-34 | 3, 7 | 18+1 | 46,XN | 46,XN | 46,XN | unaffected | 3.03 | 0.97 |
| 255 | 30-34 | 3, 5, 8 | 18+1 | 46,XN | 46,XN,t(1;4)(q41;q34) mat | 46,XN | unaffected | 1.91 | 2.09 |
| 256 | 30-34 | 9 | 16+4 | 46,XN | 46,XN | 46,XN | unaffected | 1.91 | 2.09 |
| 257 | 35-39 | 4 | 17+3 | 46,XN | 46,XN | 46,XN | unaffected | 1.96 | 2.04 |
| 258 | 35-39 | 4, 7 | 19+1 | 46,XN | 46,XN | 46,XN | unaffected | 1.98 | 2.02 |
| 259 | 30-34 | 3 | 21+1 | 46,XN | 46,XN | 46,XN | unaffected | 1.99 | 2.01 |
| 260 | 25-29 | 6 | 24+2 | 46,XN | 46,XN | 46,XN | unaffected | 1.95 | 2.05 |
| 261 | ≤24 | 6 | 16+4 | seq(X)×1,(Y)×2 | 46,X,idic(Y)(q11.2) | arr[hg19]Yq11.223(23955828-26123230)×0 arr[hg19]Yq11.223(22513726-23539614)×2 | unaffected | 1.96 | 1.04 |
| 262 | 30-34 | 7 | 29+1 | 46,XN | 46,XN | 46,XN | carrier | 0.99 | 1.01 |
| 263 | 30-34 | 7 | 16+4 | seq(21)×3 | 47,XN,+21 | arr(21)×3 | unaffected | 2.48 | 2.52 |
| 264 | 30-34 | 7 | 17+2 | 46,XN | 46,XN | 46,XN | unaffected | 1.8 | 1.2 |
| 265 | 30-34 | 9 | 17+5 | 46,XN | 46,XN | 46,XN | unaffected | 1.95 | 2.05 |
| 266 | 25-29 | 3, 7 | 18+6 | 46,XN | 46,XN | 46,XN | unaffected | 1.92 | 0.08 |
| 267 | 35-39 | 3, 4, 6 | 16+2 | 46,XN | 46,XN | 46,XN | unaffected | 1.92 | 2.08 |
| 268 | 25-29 | 6 | 18+5 | 46,XN | 46,XN | 46,XN | unaffected | 1.99 | 2.01 |
| 269 | 35-39 | 3, 4, 7 | 19 | 46,XN | 46,XN | 46,XN | unaffected | 2.01 | 1.99 |
| 270 | ≥40 | 4 | 17+6 | 46,XN | 46,XN | 46,XN | unaffected | 1.85 | 3.15 |
| 271 | 35-39 | 3, 4 | 16+6 | 46,XN | 46,XN | 46,XN | unaffected | 1.98 | 2.02 |
| 272 | 30-34 | 3, 7 | 19+3 | 46,XN | 46,XN | 46,XN | unaffected | 1.98 | 2.02 |
| 273 | ≥40 | 4 | 16 | 46,XN | 46,XN | 46,XN | unaffected | 3.08 | 0.92 |
| 274 | 30-34 | 2, 3 | 18+2 | 46,XN | 46,XN | 46,XN | unaffected | 2 | 2 |
| 275 | 30-34 | 3, 7 | 17+3 | 46,XN | 46,XN | 46,XN | unaffected | 1.93 | 2.07 |
| 276 | 35-39 | 4, 5, 7 | 19+2 | 46,XN | 46,XN | 46,XN | unaffected | 2.03 | 0.97 |
| 277 | 35-39 | 4, 5, 7 | 19+2 | 46,XN | 46,XN | 46,XN | unaffected | 2 | 2 |
| 278 | 35-39 | 3, 4, 7 | 23+3 | 46,XN | 46,XN | 46,XN | unaffected | 2 | 2 |
| 279 | 35-39 | 3, 4, 7 | 23+3 | 46,XN | 46,XN | 46,XN | unaffected | 1.99 | 2.01 |

| Clinical information | | | | low-depth GS results | Karyotype results | ECMA Results | | | |
|---|---|---|---|---|---|---|---|---|---|
| sample ID | Age (years) | Group * | Gestational age | | | Main Results | SMA screening results | *SMN1* CN | *SMN2* CN |
| 280 | 30-34 | 3, 6, 7 | 28+1 | seq[hg19]dup(2)(q36.1q37.3) chr2:g.223660000_243020000dup seq[hg19]del(15)(q26.3) chr15:g.100600000_102400000del seq[hg19]del(18)(q23) chr18:g.74240000_78020000del | 46,XN,der(15)t(2;15)(q35;q26) | arr[hg19]2q36.1q37.3(223205214-243007368)×3 arr[hg19]18q23(74216219-78015180)×1 arr[hg19]15q26.3(100601738-102400037)×1 | unaffected | 1.96 | 2.04 |
| 281 | 35-39 | 4 | 19+2 | 46,XN | 46,XN | 46,XN | unaffected | 2.06 | 1.94 |
| 282 | 35-39 | 4 | 16+5 | 46,XN | 46,XN | 46,XN | unaffected | 2 | 2 |
| 283 | 35-39 | 3, 4 | 16+4 | 46,XN | 46,XN | 46,XN | unaffected | 1.92 | 2.08 |
| 284 | 25-29 | 9 | 20+4 | 46,XN | 46,XN | 46,XN | unaffected | 1.99 | 2.01 |
| 285 | 30-34 | 3 | 16+5 | 46,XN | 46,XN | 46,XN | unaffected | 2.01 | 1.99 |
| 286 | 25-29 | 6 | 23 | 46,XN | 46,XN | 46,XN | unaffected | 2.06 | 1.94 |
| 287 | 35-39 | 4, 6 | 24+4 | 46,XN | 46,XN | 46,XN | unaffected | 3.2 | 1.8 |
| 288 | ≥40 | 3, 4 | 17+5 | 46,XN | 46,XN | 46,XN | unaffected | 1.95 | 2.05 |
| 289 | 30-34 | 3, 6 | 17 | 46,XN | 46,XN | 46,XN | unaffected | 1.97 | 2.03 |
| 290 | 35-39 | 4 | 17+5 | 46,XN | 46,XN | 46,XN | unaffected | 1.94 | 0.06 |
| 291 | 25-29 | 3, 6 | 22+4 | 46,XN | 46,XN | 46,XN | unaffected | 1.94 | 2.06 |
| 292 | ≥40 | 3, 4, 5 | 19 | 46,XN | 46,XN | 46,XN | unaffected | 2.03 | 0.97 |
| 293 | ≤24 | 7 | 18 | 46,XN | 46,XN | 46,XN | unaffected | 1.98 | 1.02 |
| 294 | 35-39 | 3, 4, 6 | 23 | 46,XN | 46,XN | 46,XN | unaffected | 1.95 | 1.05 |
| 295 | 35-39 | 3, 4 | 19+2 | 46,XN | 46,XN | 46,XN | unaffected | 1.95 | 1.05 |
| 296 | 35-39 | 4, 6 | 23+4 | 46,XN | 46,XN | 46,XN | unaffected | 1.97 | 2.03 |
| 297 | 30-34 | 9 | 23+4 | 46,XN | 46,XN | 46,XN | unaffected | 2.78 | 2.22 |
| 298 | 25-29 | 3, 6 | 24+6 | 46,XN | 46,XN | 46,XN | unaffected | 1.96 | 1.04 |
| 299 | 25-29 | 7 | 18+1 | 46,XN | 46,XN | 46,XN | unaffected | 1.99 | 2.01 |
| 300 | 35-39 | 3, 4, 7 | 18+1 | seq(21)×3 | 47,XN,+21 | arr(21)×3 | unaffected | 1.96 | 1.04 |
| 301 | 30-34 | 2, 3 | 16 | 46,XN | 46,XN | 46,XN | unaffected | 1.99 | 2.01 |
| 302 | 30-34 | 1 | 16+5 | 46,XN | 46,XN | 46,XN | unaffected | 1.99 | 2.01 |
| 303 | 30-34 | 6 | 22+4 | 46,XN | 46,XN | 46,XN | unaffected | 1.97 | 2.03 |
| 304 | 30-34 | 6 | 22+3 | 46,XN | 46,XN | 46,XN | unaffected | 1.96 | 2.04 |
| 305 | 35-39 | 3, 4, 6 | 20+2 | 46,XN | 46,XN | 46,XN | unaffected | 2 | 2 |
| 306 | 30-34 | 7 | 19+2 | 46,XN | 46,XN | 46,XN | unaffected | 3.14 | 0.86 |
| 307 | 30-34 | 3 | 18+2 | 46,XN | 46,XN,inv(17)(p13q12) | 46,XN | carrier | 1.17 | 1.83 |
| 308 | 35-39 | 4, 6 | 22+4 | 46,XN | 46,XN | 46,XN | unaffected | 1.99 | 2.01 |
| 309 | 30-34 | 7 | 20 | 46,XN | 46,XN | 46,XN | unaffected | 1.99 | 2.01 |
| 310 | 30-34 | 2, 3, 8 | 18 | 46,XN | 46,XN | 46,XN | unaffected | 2 | 2 |
| 311 | 25-29 | 3, 6 | 23+3 | 46,XN | 46,XN | 46,XN | unaffected | 1.94 | 2.06 |
| 312 | 30-34 | 3, 6 | 20+3 | 46,XN | 46,XN | 46,XN | unaffected | 2 | 2 |
| 313 | ≥40 | 3, 4 | 17 | 46,XN | 46,XN | 46,XN | unaffected | 1.94 | 2.06 |
| 314 | 30-34 | 3, 7 | 18+4 | seq(7)×3[0.2] | 47,XN,+7[3]/46,XN[17] | arr(7)×3[0.2] | unaffected | 2 | 1 |
| 315 | 30-34 | 3 | 17+5 | 46,XN | 46,XN | 46,XN | unaffected | 2 | 2 |
| 316 | ≥40 | 3, 4 | 19+5 | 46,XN | 46,XN | 46,XN | unaffected | 1.97 | 2.03 |
| 317 | 35-39 | 4, 7 | 19+2 | seq(X)×2,(Y)×1 | 47,XXY | arr(X)×2,(Y)×1 | unaffected | 1.95 | 2.05 |
| 318 | ≥40 | 3, 4 | 17+3 | 46,XN | 46,XN | 46,XN | unaffected | 1.98 | 2.02 |
| 319 | ≥40 | 3, 4 | 17+1 | 46,XN | 46,XN | 46,XN | unaffected | 2 | 2 |
| 320 | 25-29 | 7 | 23+4 | 46,XN | 46,XN | 46,XN | unaffected | 1.94 | 0.06 |
| 321 | 25-29 | 3, 5 | 16+6 | 46,XN | 46,XN | 46,XN | unaffected | 2.05 | 1.95 |
| 322 | 35-39 | 4, 6 | 23+5 | 46,XN | 46,XN | 46,XN | unaffected | 2.87 | 0.13 |
| 323 | 35-39 | 4, 6 | 23+5 | 46,XN | 46,XN | 46,XN | unaffected | 2.85 | 0.15 |
| 324 | 25-29 | 6 | 22+5 | 46,XN | 46,XN | 46,XN | unaffected | 1.94 | 2.06 |
| 325 | 25-29 | 6 | 22+5 | 46,XN | 46,XN | 46,XN | unaffected | 2.04 | 1.96 |
| 326 | 35-39 | 4 | 17+2 | seq[hg19]del(10)(q11.22) chr10:g.46960000_47720000del seq[hg19]del(10)(q11.22q11.23) chr10:g.49380000_51060000del | 46,XN | arr[hg19]10q11.22q11.23(49381635-52463140)×1 | unaffected | 2.04 | 1.96 |
| 327 | 35-39 | 3, 4 | 18 | 46,XN | 46,XN | 46,XN | unaffected | 1.89 | 0.11 |
| 328 | 25-29 | 7 | 18+2 | 46,XN | 46,XN | 46,XN | unaffected | 1.89 | 3.11 |
| 329 | 30-34 | 9 | 18+3 | 46,XN | 46,XN | 46,XN | unaffected | 1.99 | 1.01 |
| 330 | 30-34 | 9 | 22+5 | 46,XN | 46,XN | 46,XN | unaffected | 1.98 | 2.02 |
| 331 | 30-34 | 7 | 20+3 | 46,XN | 46,XN | 46,XN | unaffected | 1.94 | 2.06 |
| 332 | 35-39 | 3, 4, 6 | 17+3 | seq(18)×3 | 47,XN,+18 | arr(18)×3 | unaffected | 2.08 | 1.92 |
| 333 | 25-29 | 7 | 17+2 | 46,XN | 46,XN | 46,XN | unaffected | 1.93 | 1.07 |
| 334 | 30-34 | 3, 6 | 22+6 | 46,XN | 46,XN | 46,XN | unaffected | 2 | 2 |
| 335 | 35-39 | 4 | 18+3 | 46,XN | 46,XN | 46,XN | unaffected | 1.86 | 2.14 |
| 336 | ≥40 | 4, 8 | 17+3 | 46,XN | 46,XN | 46,XN | unaffected | 1.98 | 2.02 |
| 337 | 35-39 | 4, 7 | 19+3 | seq(21)×3 | 47,XN,+21 | arr(21)×3 | unaffected | 2.05 | 1.95 |
| 338 | 35-39 | 3, 4 | 18+2 | 46,XN | 46,XN | 46,XN | unaffected | 1.9 | 2.1 |
| 339 | 25-29 | 6 | 26+3 | 46,XN | 46,XN | 46,XN | unaffected | 2.02 | 1.98 |
| 340 | 35-39 | 3, 4 | 18+2 | 46,XN | 46,XN | 46,XN | unaffected | 1.98 | 1.02 |
| 341 | 25-29 | 6 | 22+2 | 46,XN | 46,XN | 46,XN | unaffected | 2.01 | 1.99 |
| 342 | 30-34 | 3, 6 | 17+6 | 46,XN | 46,XN | 46,XN | unaffected | 1.97 | 0.03 |
| 343 | 30-34 | 7 | 16+2 | 46,XN | 46,XN | 46,XN | unaffected | 2.01 | 1.99 |
| 344 | 35-39 | 1, 3, 4 | 17+1 | 46,XN | 46,XN | 46,XN | unaffected | 1.89 | 2.11 |
| 345 | 35-39 | 4 | 17+3 | 46,XN | 46,XN | 46,XN | unaffected | 2.01 | 1.99 |
| 346 | 35-39 | 3, 4, 6 | 26+4 | 46,XN | 46,XN | 46,XN | unaffected | 1.63 | 2.87 |
| 347 | 30-34 | 7 | 22+4 | 46,XN | 46,XN | 46,XN | unaffected | 2.01 | 1.99 |
| 348 | 25-29 | 6 | 22+5 | 46,XN | 46,XN | 46,XN | unaffected | 1.99 | 2.01 |
| 349 | ≤24 | 7 | 18+4 | 46,XN | 46,XN | 46,XN | unaffected | 1.9 | 1.1 |

| Clinical information | | | | low-depth GS results | Karyotype results | ECMA Results | | | |
|---|---|---|---|---|---|---|---|---|---|
| sample ID | Age (years) | Group * | Gestational age | | | Main Results | SMA screening results | *SMN1* CN | *SMN2* CN |
| 350 | 25-29 | 3 | 16+2 | 46,XN | 46,XN | 46,XN | unaffected | 2.03 | 1.97 |
| 351 | 35-39 | 3, 4, 6 | 17 | 46,XN | 46,XN | 46,XN | unaffected | 1.92 | 2.08 |
| 352 | 25-29 | 3, 6 | 16+3 | 46,XN | 46,XN | 46,XN | unaffected | 2.08 | 1.92 |
| 353 | 25-29 | 6 | 16+2 | 46,XN | 46,XN | 46,XN | unaffected | 1.96 | 1.04 |
| 354 | ≥40 | 4 | 20+5 | 46,XN | 46,XN | 46,XN | unaffected | 2.06 | 1.94 |
| 355 | 35-39 | 4 | 17+6 | 46,XN | 46,XN | 46,XN | unaffected | 1.94 | 1.06 |
| 356 | 35-39 | 3, 4 | 17+4 | 46,XN | 46,XN | 46,XN | unaffected | 2.01 | 0.99 |
| 357 | 25-29 | 7 | 18+6 | seq[hg19]del(2)(q13) chr2:g.111380000_113120000del | 46,XN | arr[hg19]2q13(111387676-113115598)×1 | unaffected | 1.97 | 1.03 |
| 358 | 35-39 | 3, 4, 7, 8 | 19+2 | seq[hg19]del(18)(p11.32p11.22) chr18:g.120000_8680000del | 46,XN,del(18)(p11.32p11.22) | arr[hg19]18p11.32p11.22(132649-8649262)×1 | unaffected | 1.96 | 1.04 |
| 359 | 30-34 | 3, 7 | 22+2 | 46,XN | 46,XN | 46,XN | unaffected | 2.01 | 0.99 |
| 360 | 30-34 | 7 | 17+2 | seq[hg19]dup(18)(q11.2) chr18:g.22920000_24840000dup | 46,XN | arr[hg19]18q11.2(22895523-24771938)×3 | unaffected | 1.93 | 2.07 |
| 361 | 35-39 | 3, 4 | 18+4 | 46,XN | 46,XN | 46,XN | unaffected | 1.98 | 3.02 |
| 362 | 35-39 | 3, 4, 6 | 16+4 | seq(21)×3 | 47,XN,+21 | arr(21)×3 | unaffected | 1.94 | 1.06 |
| 363 | 30-34 | 6 | 21+4 | 46,XN | 46,XN | 46,XN | unaffected | 1.98 | 2.02 |
| 364 | ≥40 | 4, 8 | 17+2 | 46,XN | 46,XN | arr[hg19]1p31.3p31.1(62753383-70917116)×2 hmz | unaffected | 2.05 | 1.95 |
| 365 | ≥40 | 4, 8 | 17+2 | 46,XN | 46,XN | arr[hg19]1p31.3p31.1(62760352-70917116)×2hmz | unaffected | 1.94 | 2.06 |
| 366 | 25-29 | 6 | 21+1 | 46,XN | 46,XN | 46,XN | unaffected | 1.94 | 1.06 |
| 367 | 25-29 | 6 | 21+1 | 46,XN | 46,XN | 46,XN | unaffected | 2.01 | 0.99 |
| 368 | 35-39 | 4 | 16+5 | 46,XN | 46,XN | 46,XN | unaffected | 1.96 | 2.04 |
| 369 | 25-29 | 7 | 17 | 46,XN | 46,XN | 46,XN | unaffected | 1.97 | 1.03 |
| 370 | 35-39 | 4 | 17+1 | 46,XN | 46,XN | 46,XN | unaffected | 1.94 | 1.06 |
| 371 | 25-29 | 9 | 17+5 | seq[hg19]dup(13)(q12.3q13.1) chr13:g.31080000_32840000dup | 46,XN | arr[hg19]13q12.3q13.1(31078840-32851814)×3 | unaffected | 1.92 | 1.08 |
| 372 | 30-34 | 6 | 24+1 | 46,XN | 46,XN | 46,XN | unaffected | 2 | 2 |
| 373 | 30-34 | 3 | 17+1 | 46,XN | 46,X,inv(Y)(p11.2q11.23) | 46,XN | unaffected | 1.99 | 2.01 |
| 374 | 30-34 | 7 | 28 | 46,XN | 46,XN | 46,XN | unaffected | 1.91 | 1.09 |
| 375 | 35-39 | 3, 4, 6 | 22 | 46,XN | 46,XN | 46,XN | unaffected | 2.27 | 2.23 |
| 376 | 35-39 | 4 | 16+5 | 46,XN | 46,XN | 46,XN | unaffected | 1.91 | 0.09 |
| 377 | 30-34 | 6 | 22+2 | 46,XN | 46,XN | 46,XN | unaffected | 1.98 | 2.02 |
| 378 | 35-39 | 3, 4 | 16+6 | 46,XN | 46,XN | 46,XN | carrier | 0.98 | 3.02 |
| 379 | ≥40 | 4 | 22 | 46,XN | 46,XN | 46,XN | unaffected | 1.99 | 1.01 |
| 380 | 25-29 | 2, 3, 6 | 23+6 | 46,XN | 46,XN | 46,XN | unaffected | 1.97 | 2.03 |
| 381 | 30-34 | 5, 8 | 16 | 46,XN | 46,XN | 46,XN | unaffected | 1.96 | 1.04 |
| 382 | 35-39 | 2, 4 | 16+4 | seq[hg19]del(X)(p21.1) chrX:g.31640000_32060000del | 46,XN | arr[hg19]Xp21.1(31643296-32056476)×0 | unaffected | 2.47 | 2.53 |
| 383 | 30-34 | 2, 3 | 17+3 | seq[hg19]del(X)(p21.1) chrX:g.32760000_32920000del | 46,XN | arr[hg19]Xp21.1(32764448-32916880)×0 | unaffected | 1.64 | 1.86 |
| 384 | 35-39 | 4 | 18+1 | 46,XN | 46,XN | arr(22)×2 hmz | unaffected | 1.92 | 1.08 |
| 385 | 35-39 | 4 | 18 | seq(X)×1[0.13] | 46,XN | 46,XN | unaffected | 1.93 | 2.07 |
| 386 | 25-29 | 3, 6 | 26+6 | 46,XN | 46,XN | 46,XN | unaffected | 1.95 | 2.05 |
| 387 | 30-34 | 7 | 18+3 | seq(X)×1[0.10] | 46,XN | 46,XN | unaffected | 1.83 | 2.17 |
| 388 | 30-34 | 1, 3 | 18+3 | 46,XN | 47，XN,+21[1]/46,XN[19] | 46,XN | unaffected | 1.57 | 2.93 |
| 389 | 25-29 | 6 | 22+4 | 46,XN | 46,XN | 46,XN | unaffected | 2 | 2 |
| 390 | 30-34 | 7 | 22+1 | 46,XN | 46,XN | 46,XN | unaffected | 2.02 | 1.98 |
| 391 | 30-34 | 3, 6 | 13 | seq[hg19]dup(1)(p21.1)chr1:103900000_106980000dup | 46,XN | 46,XN | unaffected | 1.87 | 1.13 |
| 392 | 25-29 | 3, 5 | 20+5 | 46,XN | 46,XN | 46,XN | unaffected | 1.9 | 0.1 |
| 393 | 30-34 | 6 | 22+2 | seq(Y)×1[0.5] | 45，X[14]/46,XY[16] | arr(Y)×1[0.5] | unaffected | 2.17 | 1.83 |
| 394 | 35-39 | 3, 4 | 17+3 | seq(21)×3 | 47,XN,+21 | arr(21)×3 | unaffected | 1.85 | 2.15 |
| 395 | 30-34 | 6 | 23 | seq[hg19]del(16)(p11.2)chr16:29640000_30200000del | 46,XN | arr[hg19]16p11.2(29434745-30343559)×1 | unaffected | 1.8 | 2.2 |
| 396 | 25-29 | 5, 8 | 20+3 | 46,XN | 46,XN | 46,XN | unaffected | 1.91 | 2.09 |
| 397 | ≤24 | 6 | 24+2 | seq[hg19]del(7)(q11.23)chr7:72720000_74140000del | 46,XN | arr[hg19]7q11.23(72718278-74192990)×1 | unaffected | 1.86 | 1.14 |
| 398 | 30-34 | 6 | 13+1 | seq(X)×1 | 45,X | arr(X)×1 | unaffected | 2.18 | 1.82 |
| 399 | 30-34 | 6, 7 | 19+1 | seq(13)×3 | 47，XN，+13 | arr(13)×3 | unaffected | 1.88 | 2.12 |
| 400 | 35-39 | 4 | 19 | seq(X)×1,(Y)×2 | 47,XYY | arr(X)×1,(Y)×2 | unaffected | 1.91 | 2.09 |
| 401 | 35-39 | 4, 6 | 12+6 | seq(18)×3 | 47,XN,+18 | arr(18)×3 | unaffected | 1.99 | 2.01 |
| 402 | 25-29 | 7 | 20+3 | 46,XN | 46,XN | 46,XN | unaffected | 1.98 | 2.02 |
| 403 | 35-39 | 3, 4 | 33+1 | 46,XN | 46,XN | 46,XN | unaffected | 1.59 | 1.41 |
| 404 | 25-29 | 6 | 22+3 | 46,XN | 46,XN,9ph | 46,XN | unaffected | 1.94 | 2.06 |
| 405 | ≥40 | 4 | 23+5 | 46,XN | 46,XN，9ph | 46,XN | unaffected | 1.86 | 1.14 |
| 406 | 25-29 | 3, 6 | 25+3 | seq[hg19]del(22)(q11.23-q12.1)chr22:25620000_25920000del | 46,XN | arr[hg19]22q11.23q12.1(25666567-25917661)×1 | unaffected | 1.96 | 2.04 |
| 407 | 25-29 | 3, 6 | 12+3 | 46,XN | 46,XN | 46,XN | unaffected | 1.82 | 2.18 |
| 408 | 30-34 | 6 | 14+1 | 46,XN | 46,XN | 46,XN | unaffected | 1.84 | 1.16 |

| Clinical information | | | | low-depth GS results | Karyotype results | ECMA Results | | | |
|---|---|---|---|---|---|---|---|---|---|
| sample ID | Age (years) | Group * | Gestational age | | | Main Results | SMA screening results | *SMN1* CN | *SMN2* CN |
| 409 | 30-34 | 3, 7 | 19+6 | seq[hg19]dup(22)(q11.23)chr22:23700000-24980000dup | 46,XN | arr[hg19]22q11.23(23583627-25049326)×3 | unaffected | 1.87 | 2.13 |
| 410 | 30-34 | 3 | 19+3 | 46,XN | 46,XN | 46,XN | unaffected | 1.97 | 2.03 |
| 411 | ≤24 | 6 | 23+2 | 46,XN | 46,XN | 46,XN | unaffected | 1.87 | 1.13 |
| 412 | 25-29 | 6 | 24+3 | seq[hg19]del(13)(q31.1)chr13:85040000-87600000del | 46,XN | arr[hg19]13q31.1(85031591-87635432)×1 | unaffected | 2.9 | 1.1 |
| 413 | 30-34 | 3, 6 | 23+2 | seq[hg19]dup(9)(q31.1)chr9:107020000-108100000dup | 46,XN | arr[hg19]9q31.1(107033097-108142624)×3 | unaffected | 1.89 | 1.61 |
| 414 | 25-29 | 6 | 12+5 | seq(X)×1 | 45,X | arr(X)×1 | unaffected | 1.8 | 2.2 |
| 415 | 30-34 | 7 | 16+6 | seq[hg19]del(18)(p11.32-p11.31)chr18:120000-4120000del | 46,XN | arr[hg19]18p11.32p11.31(132649-4150077)×1 | unaffected | 2.82 | 1.18 |
| 416 | 35-39 | 3, 4, 6 | 26+2 | 46,XN | 46,XN | 46,XN | unaffected | 2.42 | 2.58 |
| 417 | 25-29 | 3, 6 | 24+1 | 46,XN | 46,XN | 46,XN | unaffected | 1.86 | 2.14 |
| 418 | 30-34 | 8 | 18+3 | 46,XN | 46,XN | 46,XN | unaffected | 1.92 | 2.08 |
| 419 | 30-34 | 8 | 20+2 | seq[hg19]dup(16)(p13.11) chr16:g.15480000_16300000dup | 46,XN | arr[hg19]16p13.11p12.3(15420039-16951835)×3 | unaffected | 1.93 | 1.07 |
| 420 | 30-34 | 7 | 16+2 | 46,XN | 46,XN | arr[hg19]16q23.1q24.3(75220009-90148721)×2 hmz | unaffected | 1.87 | 0.13 |
| 421 | 35-39 | 4, 7 | 19+4 | 46,XN | 46,XN | 46,XN | unaffected | 1.92 | 1.08 |
| 422 | 35-39 | 3, 4 | 17+5 | 46,XN | 46,XN | 46,XN | unaffected | 1.92 | 1.08 |
| 423 | 30-34 | 3 | 22+5 | 46,XN | 46,XN | 46,XN | unaffected | 1.98 | 2.02 |
| 424 | 35-39 | 3, 4, 5 | 19+4 | 46,XN | 46,XN | 46,XN | unaffected | 2.88 | 0.12 |
| 425 | 35-39 | 3, 4 | 19+5 | 46,XN | 46,XN | 46,XN | unaffected | 1.9 | 1.1 |
| 426 | 25-29 | 5, 7, 8 | 18+4 | 46,XN | 46,XN | 46,XN | unaffected | 1.98 | 2.02 |
| 427 | 30-34 | 3, 8 | 16+4 | 46,XN | 46,XN | 46,XN | unaffected | 1.97 | 2.03 |
| 428 | 30-34 | 3 | 18+5 | 46,XN | 46,XN | 46,XN | unaffected | 1.94 | 2.06 |
| 429 | 30-34 | 3 | 18+5 | 46,XN | 46,XN | 46,XN | unaffected | 1.9 | 1.1 |
| 430 | 25-29 | 9 | 18 | 46,XN | 46,XN | 46,XN | unaffected | 1.9 | 1.1 |
| 431 | 30-34 | 6 | 22+2 | 46,XN | 46,XN | 46,XN | unaffected | 1.98 | 2.02 |
| 432 | 25-29 | 9 | 16+4 | 46,XN | 46,XN | 46,XN | unaffected | 1.94 | 2.06 |
| 433 | 35-39 | 4 | 20+6 | 46,XN | 46,XN | 46,XN | unaffected | 1.88 | 2.12 |
| 434 | 30-34 | 7 | 17+6 | seq(X)×2,(Y)×1 | 47,XXY | arr(X)×2,(Y)×1 | unaffected | 1.89 | 1.11 |
| 435 | 25-29 | 9 | 16 | 46,XN | 46,XN | 46,XN | unaffected | 1.98 | 2.02 |
| 436 | ≤24 | 6 | 22+6 | 46,XN | 46,XN | 46,XN | unaffected | 1.89 | 1.11 |
| 437 | 35-39 | 4 | 19+1 | 46,XN | 46,XN | 46,XN | unaffected | 1.99 | 2.01 |
| 438 | 30-34 | 3, 7 | 24+1 | seq(21)×3 | 47,XN,+21 | arr(21)×3 | unaffected | 1.91 | 1.09 |
| 439 | 25-29 | 6 | 20+1 | 46,XN | 46,XN | 46,XN | unaffected | 1.89 | 1.11 |
| 440 | 30-34 | 6 | 22+4 | 46,XN | 46,XN | 46,XN | unaffected | 1.98 | 2.02 |
| 441 | 35-39 | 3, 4 | 18+6 | 46,XN | 46,XN | 46,XN | unaffected | 2.02 | 2.98 |
| 442 | 30-34 | 6 | 18+2 | 46,XN | 46,XN | 46,XN | unaffected | 2 | 2 |
| 443 | 30-34 | 6 | 30+5 | 46,XN | 46,XN | 46,XN | unaffected | 1.62 | 1.38 |
| 444 | ≥40 | 3, 4 | 18+1 | 46,XN | 46,XN | 46,XN | unaffected | 1.86 | 1.14 |
| 445 | 30-34 | 1, 5, 8 | 18+5 | 46,XN | 46,XN | 46,XN | unaffected | 1.97 | 2.03 |
| 446 | 35-39 | 4 | 16+4 | 46,XN | 46,XN | 46,XN | unaffected | 1.96 | 2.04 |
| 447 | 30-34 | 2, 3 | 16+1 | 46,XN | 46,XN | 46,XN | unaffected | 2.87 | 1.13 |
| 448 | 35-39 | 4 | 16+4 | 46,XN | 46,XN | 46,XN | unaffected | 2.83 | 1.17 |
| 449 | 35-39 | 3, 4 | 15+5 | 46,XN | 46,XN | 46,XN | unaffected | 1.96 | 2.04 |
| 450 | 25-29 | 6 | 31+3 | 46,XN | 46,XN | 46,XN | unaffected | 1.85 | 3.15 |
| 451 | 25-29 | 1 | 18+1 | 46,XN | 46,XN | 46,XN | unaffected | 1.97 | 2.03 |
| 452 | ≥40 | 3, 4 | 17+1 | 46,XN | 46,XN | 46,XN | unaffected | 1.89 | 1.11 |
| 453 | 25-29 | 3, 6 | 24+3 | 46,XN | 46,XN | 46,XN | unaffected | 2 | 2 |
| 454 | ≥40 | 3, 4 | 17+5 | 46,XN | 46,XN | 46,XN | unaffected | 1.97 | 2.03 |
| 455 | 35-39 | 3, 4 | 17+2 | 46,XN | 46,XN | 46,XN | unaffected | 1.97 | 2.03 |
| 456 | ≥40 | 3, 4 | 19+2 | 46,XN | 46,XN | 46,XN | unaffected | 2.89 | 1.11 |
| 457 | 30-34 | 6 | 19+1 | 46,XN | 46,XN | 46,XN | unaffected | 1.89 | 2.11 |
| 458 | 30-34 | 3, 6 | 24 | 46,XN | 46,XN | 46,XN | unaffected | 1.97 | 2.03 |
| 459 | 25-29 | 7 | 27+1 | 46,XN | 46,XN | 46,XN | unaffected | 1.61 | 1.89 |
| 460 | 25-29 | 9 | 17+2 | 46,XN | 46,XN | 46,XN | unaffected | 1.91 | 1.09 |
| 461 | 35-39 | 3, 4 | 16+4 | 46,XN | 46,XN | 46,XN | unaffected | 1.93 | 3.07 |
| 462 | 30-34 | 9 | 19+3 | 46,XN | 46,XN | 46,XN | unaffected | 1.99 | 2.01 |
| 463 | 25-29 | 3, 5, 8 | 18+5 | 46,XN | 46,XN | 46,XN | unaffected | 1.99 | 2.01 |
| 464 | 35-39 | 4 | 16+4 | 46,XN | 46,XN | 46,XN | unaffected | 1.94 | 2.06 |
| 465 | 30-34 | 7 | 19 | 46,XN | 46,XN | 46,XN | unaffected | 1.96 | 2.04 |
| 466 | 30-34 | 3, 8 | 17+3 | 46,XN | 46,XN | 46,XN | unaffected | 1.99 | 2.01 |
| 467 | 30-34 | 3, 6 | 20+1 | 46,XN | 46,XN | 46,XN | unaffected | 1.98 | 2.02 |
| 468 | 25-29 | 7 | 19+6 | seq[hg19]del(15)(q11.2) chr15:g.22740000_23100000del | 46,XN | arr[hg19]15q11.1q11.2(20001226-22770992)×1 | unaffected | 2.02 | 1.98 |
| 469 | 35-39 | 4 | 16+4 | 46,XN | 46,XN | 46,XN | unaffected | 2.77 | 2.23 |
| 470 | ≤24 | 7 | 18+6 | 46,XN | 46,XN | arr[hg19]14q21.1q21.2(40332338-46217649)×2 hmz | unaffected | 1.92 | 1.08 |
| 471 | 30-34 | 6 | 22+5 | 46,XN | 46,XN | 46,XN | unaffected | 2.02 | 1.98 |
| 472 | ≤24 | 6 | 18+5 | 46,XN | 46,XN | 46,XN | unaffected | 1.99 | 2.01 |
| 473 | ≥40 | 3, 4 | 19+3 | 46,XN | 46,XN | 46,XN | unaffected | 2 | 2 |
| 474 | 35-39 | 3, 4 | 17+6 | 46,XN | 46,XN | 46,XN | unaffected | 2 | 2 |
| 475 | 35-39 | 4 | 22 | 46,XN | 46,XN | 46,XN | unaffected | 2.93 | 1.07 |
| 476 | 25-29 | 3, 7 | 18+3 | seq(21)×3 | 47,XN,+21 | arr(21)×3 | unaffected | 1.98 | 2.02 |
| 477 | 25-29 | 3 | 15+6 | 46,XN | 46,XN | 46,XN | unaffected | 1.91 | 1.09 |

| Clinical information | | | | low-depth GS results | Karyotype results | ECMA Results | | | |
|---|---|---|---|---|---|---|---|---|---|
| sample ID | Age (years) | Group * | Gestational age | | | Main Results | SMA screening results | *SMN1* CN | *SMN2* CN |
| 478 | 25-29 | 7 | 20+6 | 46,XN | 46,XN | 46,XN | unaffected | 1.99 | 2.01 |
| 479 | 30-34 | 7 | 22+2 | 46,XN | 46,XN | arr[hg19]10q22.3q23.1(78762966-83871324)×2 hmz | unaffected | 1.94 | 1.06 |
| 480 | 35-39 | 4 | 18+3 | 46,XN | 46,XN | 46,XN | unaffected | 1.98 | 2.02 |
| 481 | 35-39 | 3, 4 | 17 | 46,XN | 46,XN | 46,XN | unaffected | 1.94 | 1.06 |
| 482 | 35-39 | 3, 4 | 17+3 | 46,XN | 46,XN | 46,XN | unaffected | 1.96 | 2.04 |
| 483 | 35-39 | 3, 4 | 21 | 46,XN | 46,XN | 46,XN | unaffected | 1.98 | 2.02 |
| 484 | 35-39 | 3, 4, 5 | 17+2 | 46,XN | 46,XN,t(1;8)(q43;q22) mat | 46,XN | unaffected | 1.96 | 2.04 |
| 485 | 25-29 | 3, 8 | 17+6 | 46,XN | 46,XN | 46,XN | unaffected | 2.04 | 1.96 |
| 486 | 25-29 | 6 | 17+1 | 46,XN | 46,XN | 46,XN | unaffected | 1.96 | 1.04 |
| 487 | ≥40 | 3, 4 | 17+2 | 46,XN | 46,XN | 46,XN | unaffected | 2.03 | 1.97 |
| 488 | 30-34 | 3, 6 | 17+3 | 46,XN | 46,XN | 46,XN | unaffected | 1.94 | 1.06 |
| 489 | 30-34 | 7 | 17+3 | 46,XN | 46,XN | 46,XN | unaffected | 1.97 | 2.03 |
| 490 | 30-34 | 6 | 16+6 | 46,XN | 46,XN | 46,XN | unaffected | 2.06 | 1.94 |
| 491 | 35-39 | 3, 4, 7 | 18+5 | seq(21)×3 | 47,XN,+21 | arr(21)×3 | unaffected | 3 | 1 |
| 492 | ≥40 | 4 | 19+1 | 46,XN | 46,XN | 46,XN | unaffected | 2 | 2 |
| 493 | ≤24 | 6 | 23+6 | 46,XN | 46,XN | 46,XN | unaffected | 1.94 | 2.06 |
| 494 | 25-29 | 6 | 17+6 | 46,XN | 46,XN | 46,XN | unaffected | 1.98 | 2.02 |
| 495 | ≥40 | 3, 4 | 17+2 | 46,XN | 46,XN | 46,XN | unaffected | 1.99 | 2.01 |
| 496 | 30-34 | 3, 6 | 23 | 46,XN | 46,XN | 46,XN | unaffected | 1.96 | 2.04 |
| 497 | 30-34 | 7 | 17+4 | 46,XN | 46,XN | 46,XN | unaffected | 2.04 | 1.96 |
| 498 | 35-39 | 3, 4, 6 | 23 | 46,XN | 46,XN | 46,XN | unaffected | 1.85 | 0.15 |
| 499 | 35-39 | 3, 4 | 17 | 46,XN | 46,XN | 46,XN | unaffected | 1.99 | 2.01 |
| 500 | 30-34 | 6 | 22+3 | 46,XN | 46,XN | 46,XN | unaffected | 1.98 | 2.02 |
| 501 | 25-29 | 7 | 18+1 | seq(18)×3 | 47,XN,+18 | arr(18)×3 | unaffected | 1.93 | 2.07 |
| 502 | ≥40 | 4 | 18+2 | 46,XN | 46,XN | 46,XN | unaffected | 2.01 | 1.99 |
| 503 | 35-39 | 4 | 21+6 | 46,XN | 46,XN | 46,XN | unaffected | 1.98 | 2.02 |
| 504 | 35-39 | 3, 4, 7 | 22+4 | seq(21)×3 | 47,XN,+21 | arr(21)×3 | unaffected | 1.62 | 1.88 |
| 505 | 25-29 | 7 | 20 | 46,XN | 46,XN | 46,XN | unaffected | 1.97 | 2.03 |
| 506 | 35-39 | 4 | 17 | 46,XN | 46,XN | 46,XN | unaffected | 2.67 | 2.33 |
| 507 | 25-29 | 6 | 25+6 | 46,XN | 46,XN | 46,XN | unaffected | 1.87 | 0.13 |
| 508 | 30-34 | 5 | 13+4 | 46,XN | 46,XN | arr[hg19]19p13.11p11(18096073-31419057)×2 hmz | unaffected | 1.91 | 2.09 |
| 509 | 25-29 | 9 | 17+1 | 46,XN | 46,XN | arr[hg38]16p13.3(219828-223754)×0<br>arr[hg19]14q31.3q32.31(89463748-101613118)×2 hmz<br>arr[hg19]1q25.3q31.3(184306441-195926095)×2 hmz<br>arr[hg19]2q35q37.1(220848941-234925075)×2 hmz<br>arr[hg19]3p24.2p14.2(24314066-60342439)×2 hmz<br>arr[hg19]3q21.3q23(127484431-140074782)×2 hmz<br>arr[hg19]7p15.3p13(20912112-43406805)×2 hmz<br>arr[hg19]10q23.1(82033594-87651609)×2 hmz<br>arr[hg19]11q24.2q25(127720615-134938847)×2 hmz<br>arr[hg19]12q21.2q21.33(78114953-91779111)×2 hmz<br>arr[hg19]15q26.1q26.3(92553697-100817385)×2 hmz<br>arr[hg19]19p13.2(7235450-13350437)×2 hmz<br>arr[hg19]22q11.23q13.1(23881587-39711351)×2 hmz | unaffected | 1.87 | 0.13 |
| 510 | 30-34 | 3, 5 | 20+3 | 46,XN | 45,X,[24]/47,XXX[6] | 46,XN | unaffected | 2 | 2 |
| 511 | 30-34 | 6 | 24+2 | 46,XN | 46,XN | 46,XN | unaffected | 1.95 | 1.05 |
| 512 | 25-29 | 6 | 19+5 | 46,XN | 46,XN | 46,XN | unaffected | 1.9 | 2.1 |

*:1. Prior child with structural birth defect; 2. previous fetus or child with autosomal trisomy or sex chromosome aneuploidy; 3. Recurrent miscarriage (≥2); 4. Advanced maternal age (≥35); 5. parental carrier of a genetic disorder; 6. structural anomalies identified by ultrasonography; 7. chromosome anomalies identified by non-invasive prenatal screening; 8. validation of clinical preimplantation genetic testing (PGT); 9. Others.
46,XN: Indicates negative findings with a normal complement of 46 chromosomes, wherein 'N' represents either an X or a Y chromosome.
ECMA: expanded chromosomal microarray; low-depth GS: low-depth genome sequencing; SMA: spinal muscular atrophy.

Table S2. Screening gene list for ECMA

| Gene | Probes numbers | Locus numbers | Gene MIM | Disorder | Disorder MIM | Screening gene list during pregnancy and preconception (PMID: 34285390) | Secondary findings gene list for WES (PMID: 34012068) | Chinese newborn screening gene list (PMID: 35292922) | Fetus screening gene list (PMID: 38253798) |
|---|---|---|---|---|---|---|---|---|---|
| *ABCC8* | 782 | 738 | 600509 | Hyperinsulinism | 256450 | √ | - | √ | - |
| *ABCD1* | 1164 | 674 | 300371 | Adrenoleukodystrophy | 300100 | - | - | √ | - |
| *ACADM* | 363 | 189 | 607008 | Medium chain acyl CoA dehydrogenase deficiency | 201450 | √ | - | √ | - |
| *ACADVL* | 637 | 346 | 609575 | Very long chain acyl-CoA dehydrogenase deficiency | 201475 | √ | - | √ | - |
| *ACAT1* | 233 | 123 | 607809 | Alpha-methylacetoacetic aciduria | 203750 | √ | - | - | - |
| *APOB* | 407 | 396 | 107730 | Hypercholesterolaemia | 144010 | - | √ | - | - |
| *ARSA* | 642 | 298 | 607574 | Metachromatic leukodystrophy | 250100 | √ | - | - | - |
| *ATP7B* | 2046 | 1053 | 606882 | Wilson disease | 277900 | - | √ | √ | - |
| *BCKDHB* | 234 | 122 | 248611 | Maple syrup urine disease; type Ib | 248600 | √ | - | - | - |
| *BTD* | 554 | 279 | 609019 | Biotinidase deficiency | 253260 | √ | √ | √ | - |
| *COL1A1* | 1585 | 1310 | 120150 | Osteogenesis imperfecta III | 259420 | - | - | - | √ |
| *COL1A2* | 826 | 685 | 120160 | Osteogenesis imperfecta, type II | 166210 | - | - | - | √ |
| | | | | Osteogenesis imperfecta, type III | 259420 | - | - | - | √ |
| | | | | Osteogenesis imperfecta, type IV | 166220 | - | - | - | √ |
| *COL2A1* | 745 | 655 | 120140 | Achondrogenesis, type II or hypochondrogenesis | 200610 | - | - | - | √ |
| | | | | Avascular necrosis of the femoral head | 608805 | - | - | - | √ |
| | | | | Czech dysplasia | 609162 | - | - | - | √ |
| | | | | Epiphyseal dysplasia, multiple, with myopia and deafness | 132450 | - | - | - | √ |
| | | | | Kniest dysplasia | 156550 | - | - | - | √ |
| | | | | Legg-Calve-Perthes disease | 150600 | - | - | - | √ |
| | | | | Osteoarthritis with mild chondrodysplasia | 604864 | - | - | - | √ |
| | | | | Platyspondylic skeletal dysplasia, Torrance type | 151210 | - | - | - | √ |
| | | | | SED congenita | 183900 | - | - | - | √ |
| | | | | SMED Strudwick type | 184250 | - | - | - | √ |
| | | | | Spondyloepiphyseal dysplasia, Stanescu type | 616583 | - | - | - | √ |
| | | | | Spondyloperipheral dysplasia | 271700 | - | - | - | √ |
| | | | | Stickler sydrome, type I, nonsyndromic ocular | 609508 | - | - | - | √ |
| | | | | Stickler syndrome, type I | 108300 | - | - | - | √ |
| | | | | Vitreoretinopathy with phalangeal epiphyseal dysplasia | 619248 | - | - | - | √ |
| *CPT1A* | 87 | 53 | 600528 | Carnitine palmitoyltransferase 1 deficiency | 255120 | - | - | √ | - |
| *CPT2* | 140 | 127 | 600650 | CPT II deficiency, infantile | 600649 | √ | - | - | - |
| | | | | CPT II deficiency, lethal neonatal | 608836 | √ | - | - | - |
| *ETFA* | 59 | 33 | 608053 | Glutaric acidaemia 2a | 231680 | - | - | √ | - |
| *ETFDH* | 434 | 226 | 231675 | Glutaric acidaemia 2c | 231680 | - | - | √ | - |
| *FAH* | 180 | 104 | 613871 | Tyrosinaemia 1 | 276700 | √ | - | - | - |
| *FGF23* | 42 | 23 | 605380 | Rickets, hypophosphataemic, autosomal dominant | 193100 | - | - | √ | - |
| *FGFR1* | 299 | 286 | 136350 | Hartsfield syndrome | 615465 | - | - | - | √ |
| | | | | Hypogonadotropic hypogonadism 2 with or without anosmia | 147950 | - | - | - | √ |
| | | | | Jackson-Weiss syndrome | 123150 | - | - | - | √ |
| | | | | Osteoglophonic dysplasia | 166250 | - | - | - | √ |
| | | | | Pfeiffer syndrome | 101600 | - | - | - | √ |
| | | | | Trigonocephaly 1 | 190440 | - | - | - | √ |
| *FGFR2* | 171 | 143 | 176943 | Antley-Bixler syndrome without genital anomalies or disordered steroidogenesis | 207410 | - | - | - | √ |
| | | | | Apert syndrome | 101200 | - | - | - | √ |
| | | | | Beare-Stevenson cutis gyrata syndrome | 123790 | - | - | - | √ |
| | | | | Bent bone dysplasia syndrome | 614592 | - | - | - | √ |
| | | | | Craniofacial-skeletal-dermatologic dysplasia | 101600 | - | - | - | √ |
| | | | | Crouzon syndrome | 123500 | - | - | - | √ |
| | | | | Gastric cancer, somatic | 613659 | - | - | - | √ |
| | | | | Jackson-Weiss syndrome | 123150 | - | - | - | √ |
| | | | | LADD syndrome | 149730 | - | - | - | √ |
| | | | | Pfeiffer syndrome | 101600 | - | - | - | √ |
| | | | | Saethre-Chotzen syndrome | 101400 | - | - | - | √ |
| | | | | Scaphocephaly, maxillary retrusion, and mental retardation | 609579 | - | - | - | √ |
| *FGFR3* | 134 | 75 | 134934 | Achondroplasia | 100800 | - | - | - | √ |
| *G6PC* | 242 | 129 | 613742 | Glycogen storage disease 1a | 232200 | √ | - | - | - |
| *GAA* | 1178 | 657 | 606800 | Glycogen storage disease 2 | 232300 | √ | √ | - | - |
| *GALT* | 715 | 378 | 606999 | Galactosaemia | 230400 | √ | - | - | - |
| *GBA* | 877 | 477 | 606463 | Gaucher disease, perinatal lethal | 608013 | √ | - | - | - |
| | | | | Gaucher disease, type I | 230800 | √ | - | √ | - |
| | | | | Gaucher disease, type II | 230900 | √ | - | - | - |
| | | | | Gaucher disease, type III | 231000 | √ | - | - | - |
| | | | | Gaucher disease, type IIIC | 231005 | √ | - | - | - |
| *GBE1* | 154 | 82 | 607839 | Glycogen storage disease 4 | 232500 | √ | - | - | - |
| *GCDH* | 512 | 266 | 608801 | Glutaric acidaemia 1 | 231670 | - | - | √ | - |
| *GCH1* | 507 | 263 | 600225 | Hyperphenylalaninemia; BH4-deficient; B | 233910 | - | - | √ | - |
| *GJB2* | 1052 | 434 | 121011 | Deafness, autosomal recessive 1A | 220290 | √ | - | √ | - |
| | | | | Deafness, autosomal dominant 3A | 601544 | √ | - | - | - |

| Gene | Probes numbers | Locus numbers | Gene MIM | Disorder | Disorder MIM | Screening gene list during pregnancy and preconception (PMID: 34285390) | Secondary findings gene list for WES (PMID: 34012068) | Chinese newborn screening gene list (PMID: 35292922) | Fetus screening gene list (PMID: 38253798) |
|---|---|---|---|---|---|---|---|---|---|
| *GLA* | 2218 | 1054 | 300644 | Fabry disease | 301500 | - | √ | √ | - |
| *GNPTAB* | 536 | 289 | 607840 | Mucolipidosis II | 252500 | √ | - | - | - |
| *HBB* | 2696 | 916 | 141900 | Delta-beta thalassemia | 141749 | √ | - | - | - |
| *HEXA* | 536 | 248 | 606869 | Tay-Sachs disease | 272800 | √ | - | - | - |
| *HLCS* | 88 | 48 | 609018 | Holocarboxylase synthetase deficiency | 253270 | - | - | √ | - |
| *IDUA* | 494 | 297 | 252800 | Mucopolysaccharidosis Ih | 607014 | √ | - | - | - |
| | | | | Mucopolysaccharidosis Ih/s | 607015 | √ | - | - | - |
| | | | | Mucopolysaccharidosis Is | 607016 | √ | - | - | - |
| *IFITM5* | 7 | 5 | 614757 | Osteogenesis imperfecta 5 | 610967 | - | - | - | √ |
| *IVD* | 187 | 107 | 607036 | Isovaleric acidaemia | 243500 | - | - | √ | - |
| *LDLR* | 2062 | 1838 | 606945 | Hypercholesterolaemia | 143890 | - | √ | - | - |
| *MCCC2* | 258 | 128 | 609014 | 3-Methylcrotonyl-CoA carboxylase 2 deficiency | 210210 | √ | - | √ | - |
| *MCOLN1* | 51 | 34 | 605248 | Mucolipidosis IV | 252650 | √ | - | - | - |
| *MMACHC* | 276 | 122 | 609831 | Methylmalonic aciduria;cblC type | 277400 | √ | - | - | - |
| *MMUT* | 894 | 419 | 609058 | Methylmalonic aciduria;mut(0)type | 251000 | √ | - | - | - |
| *OTC* | 973 | 498 | 300461 | Ornithine transcarbamylase deficiency | 311250 | - | √ | √ | - |
| *PAH* | 2165 | 1055 | 612349 | Phenylketonuria | 261600 | √ | - | - | - |
| *PCCA* | 270 | 143 | 232000 | Propionic acidaemia | 606054 | - | - | √ | - |
| *PCCB* | 276 | 144 | 232050 | Propionic acidaemia | 606054 | - | - | √ | - |
| *POLG* | 738 | 365 | 174763 | Mitochondrial DNA depletion syndrome 4A | 203700 | √ | - | - | - |
| | | | | Mitochondrial DNA depletion syndrome 4B | 613662 | √ | - | - | - |
| | | | | Mitochondrial recessive ataxia syndrome | 607459 | √ | - | - | - |
| *SLC22A5* | 292 | 164 | 603377 | Carnitine deficiency, systemic primary | 212140 | - | - | √ | - |
| *SLC25A20* | 76 | 42 | 613698 | Carnitine-acylcarnitine translocase deficiency | 212138 | - | - | √ | - |
| *SLC26A4* | 1465 | 667 | 605646 | Deafness, autosomal recessive 4, with enlarged vestibular aqueduct Pendred syndrome | 600791 | √ | - | - | - |
| *SLC37A4* | 273 | 129 | 602671 | Glycogen storage disease 1b | 232220 | √ | - | - | - |
| *SMN1* | 172 | 93 | 600354 | Spinal muscular atrophy-3 | 253400 | - | - | √ | - |
| *SMPD1* | 443 | 261 | 607608 | Niemann-Pick disease, type A | 257200 | √ | - | √ | - |
| | | | | Niemann-Pick disease, type B | 607616 | √ | - | - | - |
| *TAT* | 62 | 34 | 613018 | Tyrosinaemia 2 | 276600 | - | - | √ | - |

ECMA: expanded chromosomal microarray; MIM: number of Online Mendelian Inheritance in Man database; WES: whole exome sequecing.

**Table S3. Summary of 31 aneuploidies detected by ECMA, low-depth GS, and karyotype analysis**

| Characteristics | Summary (n=31) | Trisomy 21 (n=18) | Trisomy 18 (n=3) | Klinefelter Syndrome (n=3) | Turner Syndrome (n=3) | Trisomy X (n=2) | Trisomy 13 (n=1) | Jacobs Syndrome (n=1) |
|---|---|---|---|---|---|---|---|---|
| Mean maternal age—year (range) | 33.9 (22-43) | 36.7 (26-43) | 35.3 (29-39) | 35 (32-37) | 27.3 (22-32) | 28.5 (27-30) | 31 | 37 |
| Median maternal age—year (range) | 35 (22-43) | 35 (26-43) | 38 (29-39) | 36 (32-37) | 28 (22-32) | - | - | - |
| Mothers ≥35 years old—no. (%) | 16 (51.6) | 11 (61.1) | 2 (66.7) | 2 (66.7) | 0 (0) | 0 (0) | 0 (0) | 1 (100.0) |

### Table S4. 29 Multigene CNVs detected by ECMA and low-depth GS

| Sample ID | Low depth GS results | ECMA results | Pathogenicity | Karyotype results |
|---|---|---|---|---|
| 4 | seq[hg19]dup(15)(q11.2q13.1) chr15:g.23620000_28420000dup | arr[hg19]15q11.2q13.1(23616115-28555716)×3 | LP | 46,XN |
| 59 | seq[hg19]dup(1)(q44) chr1:g.246220000_247500000dup | arr[hg19]1q44(246217629-247516609)×3 | VUS | 46,XN |
| 63 | seq[hg19]del(4)(q22.1q22.3) chr4:g.93300000_96960000del | arr[hg19]4q22.1q22.3(93296704-97008926)×1 | VUS | 46,XN |
| 82 | seq[hg19]dup(11)(q21q22.1) chr11:g.95740000_98300000dup | arr[hg19]11q21q22.1(95726001-98289956)×3 | VUS | 46,XN,t(4;8)(q35;q13)mat |
| 135 | seq[hg19]del(X)(p22.12p22.11) chrX:g.20500000_23860000del | arr[hg19]Xp22.12p22.11(20536377-23832515)×1 | P | 46,XN |
| 138 | seq[hg19]dup(13)(q21.31q34) chr13:g.64440000_115100000dup | arr[hg19]13q21.31q34(64407224-114800246)×3 | P | 47,XN,+der(13)del(13)(q11q21.3) |
| 179 | seq[hg19]del(12)(p13.33p13.31) chr12:g.160000_5880000del seq[hg19]dup(15)(q22.31q26.3) chr15:g.63720000_102400000dup | arr[hg19]12p13.33p13.31(83711-5897339)×1 arr[hg19]15q22.32q26.1(67202258-89337616)×3 | P; P | 46，XN,add(12)(p13)[25]/46, XN[5] |
| 184 | seq[hg19]dup(22)(q11.21q11.23) chr22:g.18880000_25180000dup | arr[hg19]22q11.21q11.23(18614445-25127865)×3 | P | 46,XN |
| 188 | seq[hg19]del(15)(q13.3) chr15:g.32020000_32520000del | arr[hg19]15q13.3(32014503-32444185)×1 | P | 46,XN |
| 207 | seq[hg19]del(X)(p22.31) chrX:g.6440000_8140000del | arr[hg19]Xp22.31(6454813-8126718)×0 | P | 46,XN |
| 228 | seq[hg19]del(X)(q24) chrX:g.118960000_119100000del | arr[hg19]Xq24(118940712-119104408)×1 | P | 46,XN |
| 250 | seq[hg19]del(11)(p14.3) chr11:g.23320000_24680000del | arr[hg19]11p14.3(23337011-24681532)×1 | VUS | 46,XN |
| 261 | seq(X)×1,(Y)×2 | arr[hg19]Yq11.223(23955828-26123230)×0 arr[hg19]Yq11.223(22513726-23539614)×2 | P; P | 46,X,idic(Y)(q11.2) |
| 280 | seq[hg19]dup(2)(q36.1q37.3) chr2:g.223660000_243020000dup seq[hg19]del(15)(q26.3) chr15:g.100600000_102400000del seq[hg19]del(18)(q23) chr18:g.74240000_78020000del | arr[hg19]2q36.1q37.3(223205214-243007368)×3 arr[hg19]18q23(74216219-78015180)×1 arr[hg19]15q26.3(100601738-102400037)×1 | LP; VUS; VUS | 46,XN,der(15)t(2;15)(q35;q26) |
| 326 | seq[hg19]del(10)(q11.22) chr10:g.46960000_47720000del seq[hg19]del(10)(q11.22q11.23) chr10:g.49380000_51060000del | arr[hg19]10q11.22q11.23(49381635-52463140)×1 | VUS; | 46,XN |
| 357 | seq[hg19]del(2)(q13) chr2:g.111380000_113120000del | arr[hg19]2q13(111387676-113115598)×1 | LP | 46,XN |
| 358 | seq[hg19]del(18)(p11.32p11.22) chr18:g.120000_8680000del | arr[hg19]18p11.32p11.22(132649-8649262)×1 | P | 46,XN,del(18)(p11.32p11.22) |
| 360 | seq[hg19]dup(18)(q11.2) chr18:g.22920000_24840000dup | arr[hg19]18q11.2(22895523-24771938)×3 | VUS | 46,XN |
| 371 | seq[hg19]dup(13)(q12.3q13.1) chr13:g.31080000_32840000dup | arr[hg19]13q12.3q13.1(31078840-32851814)×3 | VUS | 46,XN |
| 391 | seq[hg19]dup(1)(p21.1)chr1:103900000_106980000dup | arr[hg19]1p21.1(103926241-106985964)×3 | VUS | 46,XN |
| 395 | seq[hg19]del(16)(p11.2)chr16:29640000_30200000del | arr[hg19]16p11.2(29434745-30343559)×1 | P | 46,XN |
| 397 | seq[hg19]del(7)(q11.23)chr7:72720000_74140000del | arr[hg19]7q11.23(72718278-74192990)×1 | P | 46,XN |
| 406 | seq[hg19]del(22)(q11.23-q12.1)chr22:25620000_25920000de | arr[hg19]22q11.23q12.1(25666567-25917661)×1 | VUS | 46,XN |
| 409 | seq[hg19]dup(22)(q11.23)chr22:23700000-24980000dup | arr[hg19]22q11.23(23583627-25049326)×3 | VUS | 46,XN |
| 412 | seq[hg19]del(13)(q31.1)chr13:85040000-87600000del | arr[hg19]13q31.1(85031591-87635432)×1 | VUS | 46,XN |
| 413 | seq[hg19]dup(9)(q31.1)chr9:107020000-108100000dup | arr[hg19]9q31.1(107033097-108142624)×3 | VUS | 46,XN |
| 415 | seq[hg19]del(18)(p11.32-p11.31)chr18:120000-4120000del | arr[hg19]18p11.32p11.31(132649-4150077)×1 | P | 46,XN |
| 419 | seq[hg19]dup(16)(p13.11) chr16:g.15480000_16300000dup | arr[hg19]16p13.11p12.3(15420039-16951835)×3 | LP | 46,XN |
| 468 | seq[hg19]del(15)(q11.2) chr15:g.22740000_23100000del | arr[hg19]15q11.1q11.2(20001226-22770992)×1 | LP | 46,XN |

46,XN: Indicates negative findings with a normal complement of 46 chromosomes, wherein 'N' represents either an X or a Y chromosome.
CNVs:copy number variations; ECMA: expanded chromosomal microarray; low-depth GS: low-depth genome sequencing; P: pathogenic; LP: like pathogenic; VUS: variant of uncertain significance.

**Table S5. 2 *DMD* exonic CNVs detected by ECMA and low-depth GS**

| Sample ID | ECMA results | | low-depth GS results | | Karyotype results | Deleted exons of *DMD* |
|---|---|---|---|---|---|---|
| 382 | arr[hg19]Xp21.1(31643296-32056476)×0 | 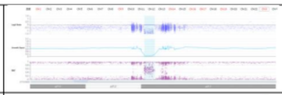 | seq[hg19]del(X)(p21.1) chrX:g.31640000_32060000del | 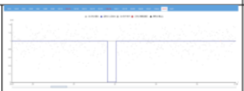 | 46,XN | 3-7 |
| 383 | arr[hg19]Xp21.1(32764448-32916880)×0 | 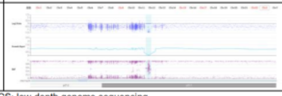 | seq[hg19]del(X)(p21.1) chrX:g.32760000_32920000del | 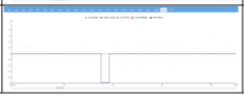 | 46,XN | 45-55 |

ECMA: expanded chromosomal microarray; low-depth GS: low-depth genome sequencing.

**Table S6. 111 SNVs identified by ECMA and validated by Sanger sequencing in 511 fetuses**

| Sample ID | ECMA results | | Sanger sequencing results | | | Pathogenicity classification |
|---|---|---|---|---|---|---|
| | Locus | Genotypic status | Genotypic status of ECMA locus | Valid Locus | Genotypic status of valid Locus | |
| 8 | NM_000441(*SLC26A4*):c.A1124G(p.Y375C) | heterozygote | heterozygote | NM_000441(*SLC26A4*):c.1124A>G(p.Y375C) | heterozygote | VUS |
| 8 | NM_000441(*SLC26A4*):c.G1409A(p.R470H) | heterozygote | heterozygote | NM_000441(*SLC26A4*):c.1409G>A(p.R470H) | heterozygote | VUS |
| 10 | NM_000441(*SLC26A4*):c.A2168G(p.H723R) | heterozygote | heterozygote | NM_000441(*SLC26A4*):c.2168A>G(p.H723R) | heterozygote | P |
| 17 | NM_000089(*COL1A2*):c.G2933A(p.R978H) | heterozygote | heterozygote | NM_000089(*COL1A2*):c.2933G>A(p.R978H) | heterozygote | VUS |
| 27 | NM_000053(*ATP7B*):c.C3008T(p.A1003V) | heterozygote | wild type | NM_000053(*ATP7B*):c.3009G>A(p.Ala1003=) | heterozygote | B |
| 28 | NM_000518(*HBB*):c.A247C(p.K83Q) | heterozygote | heterozygote | NM_000518(*HBB*):c.246C>A(p.Leu82=) | heterozygote | LB |
| 32 | NM_000384(*APOB*):c.G288T(p.Q96H) | heterozygote | heterozygote | NM_000384(*APOB*):c.288G>T(p.Q96H) | heterozygote | LB |
| 46 | NM_000142(*FGFR3*):c.G1138A(p.G380R) | heterozygote | heterozygote | NM_000142(*FGFR3*):c.1138G>A(p.G380R) | heterozygote | P |
| 80 | NM_000277(*PAH*):c.G158A(p.R53H) | heterozygote | heterozygote | NM_000277(*PAH*):c.158G>A(p.R53H) | heterozygote | VUS |
| 82 | NM_000531(*OTC*):c.C621A(p.S207R) | heterozygote | heterozygote | NM_000531(*OTC*):c.621C>T(p.Ser207=) | heterozygote | B |
| 83 | NM_000527(*LDLR*):c.C2538G(p.S846R) | heterozygote | heterozygote | NM_000527(*LDLR*):c.2538C>G(p.S846R) | heterozygote | VUS |
| 95 | NM_000277(*PAH*):c.941delC(p.P314fs) | heterozygote | wild type | NM_000277(*PAH*):c.940C>A(p.Pro314Thr) | heterozygote | P |
| 100 | NM_000053(*ATP7B*):c.3892_3894del(p.1298_1298del | heterozygote | wild type | NM_000053(*ATP7B*):c.3886G>A(p.Asp1296Asn) | heterozygote | LP |
| 109 | NM_000441(*SLC26A4*):c.C1229A(p.T410K) | heterozygote | heterozygote | NM_000441(*SLC26A4*):c.1229C>T(p.Thr410Met) | heterozygote | P |
| 116 | NM_000384(*APOB*):c.C10700T(p.T3567M) | heterozygote | heterozygote | NM_000384(*APOB*):c.10700C>T(p.T3567M) | heterozygote | VUS |
| 116 | NM_000088(*COL1A1*):c.G1522A(p.A508T) | heterozygote | heterozygote | NM_000088(*COL1A1*):c.1522G>A(p.A508T) | heterozygote | LB |
| 122 | NM_000088(*COL1A1*):chr17:50192028AG>AT | heterozygote | wild type | - | wild type | - |
| 126 | NM_000053(*ATP7B*):c.3892_3894del(p.1298_1298del | heterozygote | wild type | NM_000053(*ATP7B*):c.3886G>A(p.Asp1296Asn) | heterozygote | LP |
| 138 | NM_000255(*MMUT*):c.G1663A(p.A555T) | heterozygote | heterozygote | NM_000255(*MMUT*):c.1663G>A(p.A555T) | heterozygote | LP |
| 138 | NM_000255(*MMUT*):c.1332+1G>- | heterozygote | wild type | - | wild type | - |
| 146 | NM_000384(*APOB*):c.G288T(p.Q96H) | heterozygote | heterozygote | NM_000384(*APOB*):c.288G>T(p.Q96H) | heterozygote | LB |
| 151 | NM_001005741(*GBA*):c.G721A(p.G241R) | heterozygote | wild type | - | wild type | - |
| 154 | NM_001025295(*IFITM5*):c.C119T(p.S40L) | heterozygote | wild type | NM_001025295(*IFITM5*):c.120G>T(p.Ser40=) | heterozygote | B |
| 181 | NM_001025295(*IFITM5*):c.C119T(p.S40L) | heterozygote | wild type | NM_001025295(*IFITM5*):c.120G>T(p.Ser40=) | heterozygote | B |
| 181 | NM_000152(*GAA*):c.1441delT(p.W481fs) | heterozygote | wild type | - | wild type | - |
| 181 | NM_000152(*GAA*):c.G1912T(p.G638W) | heterozygote | wild type | - | wild type | - |
| 188 | NM_000384(*APOB*):c.A3607G(p.S1203G) | heterozygote | heterozygote | NM_000384(*APOB*):c.3607A>G(p.S1203G) | heterozygote | LB |
| 193 | NM_000527(*LDLR*):c.C858A(p.S286R) | heterozygote | heterozygote | NM_000527(*LDLR*):c.858C>T(p.Ser286=) | homozygote | LB |
| 210 | NM_000441(*SLC26A4*):c.C1983A(p.D661E) | heterozygote | heterozygote | NM_000441(*SLC26A4*):c.1983C>A(p.D661E) | heterozygote | LP |

| Sample ID | ECMA results | | Sanger sequencing results | | | Pathogenicity classification |
|---|---|---|---|---|---|---|
| | Locus | Genotypic status | Genotypic status of ECMA locus | Valid Locus | Genotypic status of valid Locus | |
| 212 | NM_000155(*GALT*):chr9:34646576CAGT>- | heterozygote | heterozygote | NM_000155.4(*GALT*):c.-119_-116del | heterozygote | VUS |
| 213 | NM_000277(*PAH*):c.842+2T>A | heterozygote | heterozygote | NM_000277(*PAH*):c.842+2T>A | heterozygote | P |
| 218 | NM_000152(*GAA*):c.G2237C(p.W746S) | heterozygote | heterozygote | NM_000152(*GAA*):c.2237G>C(p.W746S) | heterozygote | P |
| 220 | NM_001164277(*SLC37A4*):c.C572G(p.P191R) | heterozygote | heterozygote | NM_001164277(*SLC37A4*):c.572C>T(p.Pro191Leu) | heterozygote | P |
| 229 | NM_000277(*PAH*):c.1024delG(p.A342fs) | heterozygote | wild type | - | wild type | - |
| 232 | NM_001005741(*GBA*):c.A928G(p.S310G) | heterozygote | wild type | - | wild type | - |
| 232 | NM_001005741(*GBA*):c.G721A(p.G241R) | heterozygote | wild type | - | wild type | - |
| 243 | NM_000152(*GAA*):c.1580_1581del(p.R527fs) | homozygote | homozygote | NM_000152(*GAA*):c.1581G>A(p.Arg527=) | heterozygote | B |
| 246 | NM_000441(*SLC26A4*):c.G697C(p.V233L) | heterozygote | heterozygote | NM_000441(*SLC26A4*):c.697G>C(p.V233L) | heterozygote | LP |
| 246 | NM_000441(*SLC26A4*):c.681_697del(p.A227fs) | heterozygote | wild type | NM_000441(*SLC26A4*):c.680C>G(p.Ala227Gly) | heterozygote | LP |
| 247 | NM_000527(*LDLR*):c.C1056G(p.C352W) | heterozygote | heterozygote | NM_000527(*LDLR*):c.1056C>T(p.Cys352=) | heterozygote | B |
| 248 | NM_023110(*FGFR1*):c.C320A(p.S107X) | heterozygote | heterozygote | NM_023110(*FGFR1*):c.320C>T(p.Ser107Leu) | homozygote | B |
| 249 | NM_000137(*FAH*):c.C1056A(p.S352R) | homozygote | homozygote | NM_000137(*FAH*):c.1056C>T(p.Ser352=) | heterozygote | B |
| 250 | NM_000089(*COL1A2*):c.C4095A(p.F1365L) | heterozygote | heterozygote | NM_000089(*COL1A2*):c.4095C>A(p.F1365L) | heterozygote | VUS |
| 264 | NM_000518(*HBB*):c.*110T>A | heterozygote | wild type | - | wild type | - |
| 265 | NM_000088(*COL1A1*):c.G3766A(p.A1256T) | heterozygote | heterozygote | NM_000088(*COL1A1*):c.3766G>A(p.A1256T) | heterozygote | LB |
| 266 | NM_000384(*APOB*):c.A3607G(p.S1203G) | heterozygote | heterozygote | NM_000384(*APOB*):c.3607A>G(p.S1203G) | heterozygote | LB |
| 314 | NM_000169(*GLA*):c.G128T(p.G43V) | homozygote | wild type | - | wild type | - |
| 322 | NM_000518(*HBB*):c.27dupG(p.S10fs) | heterozygote | wild type | NM_000518.5(*HBB*):c.25_26insG(p.Lys9ArgfsTer15) | heterozygote | LP |
| 322 | NM_000518(*HBB*):c.26_27insAGAA(p.K9fs) | heterozygote | heterozygote | NM_000518.5(*HBB*):c.25_26insG(p.Lys9ArgfsTer15) | heterozygote | LP |
| 322 | NM_000518(*HBB*):c.25_26del(p.K9fs) | heterozygote | wild type | NM_000518.5(*HBB*):c.25_26insG(p.Lys9ArgfsTer15) | homozygote | LP |
| 322 | NM_000053(*ATP7B*):chr13:51968441->A | heterozygote | heterozygote | NM_000053.4(*ATP7B*):c.1707+2dup | heterozygote | VUS |
| 322 | NM_000089(*COL1A2*):c.G1252A(p.G418S) | heterozygote | wild type | - | wild type | - |
| 323 | NM_000518(*HBB*):c.26_27insAGAA(p.K9fs) | heterozygote | heterozygote | NM_000518.5(*HBB*):c.25_26insG(p.Lys9ArgfsTer15) | homozygote | LP |
| 323 | NM_000053(*ATP7B*):chr13:51968439C>T | heterozygote | wild type | NM_000053.4(*ATP7B*):c.1707+2dup | heterozygote | VUS |
| 324 | NM_000527(*LDLR*):c.C81A(p.C27X) | heterozygote | heterozygote | NM_000527(*LDLR*):c.81C>T(p.Cys27=) | heterozygote | B |
| 324 | NM_003060(*SLC22A5*):c.C428T(p.P143L) | homozygote | heterozygote | NM_003060(*SLC22A5*):c.428C>T(p.P143L) | heterozygote | P |
| 325 | NM_000527(*LDLR*):c.C81A(p.C27X) | heterozygote | heterozygote | NM_000527(*LDLR*):c.81C>T(p.Cys27=) | heterozygote | B |
| 325 | NM_003060(*SLC22A5*):c.C428T(p.P143L) | homozygote | heterozygote | NM_003060(*SLC22A5*):c.428C>T(p.P143L) | heterozygote | P |
| 326 | NM_000137(*FAH*):c.C1056A(p.S352R) | homozygote | homozygote | NM_000137(*FAH*):c.1056C>T(p.Ser352=) | heterozygote | B |

| Sample ID | ECMA results | | Sanger sequencing results | | | Pathogenicity classification |
|---|---|---|---|---|---|---|
| | Locus | Genotypic status | Genotypic status of ECMA locus | Valid Locus | Genotypic status of valid Locus | |
| 327 | NM_000089(*COL1A2*):c.C3794G(p.S1265C) | heterozygote | heterozygote | NM_000089(*COL1A2*):c.3794C>G(p.S1265C) | heterozygote | LP |
| 330 | NM_000137(*FAH*):c.C1056A(p.S352R) | homozygote | homozygote | NM_000137(*FAH*):c.1056C>T(p.Ser352=) | heterozygote | B |
| 341 | NM_000088(*COL1A1*):c.G1714C(p.G572R) | homozygote | wild type | - | wild type | - |
| 341 | NM_000531(*OTC*):c.C419A(p.A140D) | heterozygote | heterozygote | NM_000531(*OTC*):c.419C>T(p.Ala140Val) | heterozygote | LP |
| 342 | NM_000384(*APOB*):c.3264_3267del(p.T1088fs) | heterozygote | heterozygote | NM_000384(*APOB*):c.3264G>A(p.Thr1088=) | heterozygote | LB |
| 344 | NM_000518(*HBB*):c.C309A(p.N103K) | heterozygote | wild type | - | wild type | - |
| 344 | NM_000282(*PCCA*):c.G1676T(p.W559L) | homozygote | homozygote | NM_000282(*PCCA*):c.1676G>T(p.W559L) | homozygote | B |
| 347 | NM_000155(*GALT*):chr9:34646576 CAGT>- | heterozygote | heterozygote | NM_000155.4(*GALT*):c.-119_-116del | heterozygote | VUS |
| 356 | NM_001025295(*IFITM5*):c.C119T(p.S40L) | heterozygote | wild type | NM_001025295(*IFITM5*):c.120G>T(p.Ser40=) | heterozygote | B |
| 356 | NM_000053(*ATP7B*):c.G2333T(p.R778L) | heterozygote | heterozygote | NM_000053(*ATP7B*):c.2333G>T(p.R778L) | heterozygote | P |
| 362 | NM_000527(*LDLR*):c.G1195T(p.A399S) | heterozygote | wild type | NM_000527(*LDLR*):c.1194C>T(p.Ile398=) | neighboring mutati | B |
| 364 | NM_000527(*LDLR*):c.*2196_*2199delTATA | homozygote | homozygote | NM_000527(*LDLR*):c.*2196_*2199delTATA | homozygote | VUS |
| 369 | NM_000277(*PAH*):c.G158A(p.R53H) | heterozygote | heterozygote | NM_000277(*PAH*):c.158G>A(p.R53H) | heterozygote | VUS |
| 371 | NM_000033(*ABCD1*):c.T1673C(p.I558T) | homozygote | wild type | - | wild type | - |
| 373 | NM_000277(*PAH*):c.G158A(p.R53H) | heterozygote | heterozygote | NM_000277(*PAH*):c.158G>A(p.R53H) | heterozygote | VUS |
| 418 | NM_000018(*ACADVL*):c.1843C>T(p.R615X) | heterozygote | wild type | NM_000018(*ACADVL*):c.1844G>T(p.Arg615Leu) | heterozygote | VUS |
| 422 | NM_000018(*ACADVL*):c.776_777insCA(p.F259fs) | heterozygote | wild type | NM_000018(*ACADVL*):c.779C>T( p.Thr260Met) | heterozygote | P |
| 424 | NM_000155(*GALT*):c.652delC(p.L218X) | heterozygote | heterozygote | NM_000155(*GALT*):c.652C>T(p.L218X) | heterozygote | B |
| 429 | NM_000088(*COL1A1*):c.3766G>A(p.A1256T) | heterozygote | heterozygote | NM_000088(*COL1A1*):c.3766G>A(p.A1256T) | heterozygote | LB |
| 430 | NM_000277(*PAH*):c.157C>T(p.R53C) | heterozygote | wild type | NM_000277(*PAH*):c.158G>A(p.Arg53His) | heterozygote | VUS |
| 437 | NM_000384(*APOB*):c.3607A>G(p.S1203G) | heterozygote | heterozygote | NM_000384(*APOB*):c.3607A>G(p.S1203G) | heterozygote | LB |
| 439 | NM_000033(*ABCD1*):c.1537A>C(p.K513Q) | homozygote | wild type | - | wild type | - |
| 440 | NM_000277(*PAH*):c.697T>A(p.F233I) | homozygote | wild type | - | wild type | - |
| 440 | NM_003060(*SLC22A5*):c.1196G>A(p.R399Q) | heterozygote | heterozygote | NM_003060(*SLC22A5*):c.1196G>A(p.R399Q) | heterozygote | P |
| 444 | NM_000384(*APOB*):c.10579C>T(p.R3527W) | heterozygote | heterozygote | NM_000384(*APOB*):c.10579C>T(p.R3527W) | homozygote | LP |
| 445 | NM_023110(*FGFR1*):c.1854G>T(p.K618N) | heterozygote | wild type | - | wild type | - |
| 450 | NM_000527(*LDLR*):c.1056C>A(p.C352X) | heterozygote | heterozygote | NM_000527(*LDLR*):c.1056C>T(p.Cys352=) | heterozygote | B |
| 452 | NM_000384(*APOB*):c.3607A>G(p.S1203G) | heterozygote | heterozygote | NM_000384(*APOB*):c.3607A>G(p.S1203G) | heterozygote | LB |
| 452 | NM_000089(*COL1A2*):c.3124G>A(p.G1042S) | heterozygote | wild type | - | wild type | - |
| 458 | NM_000441(*SLC26A4*):c.1983C>A(p.D661E) | heterozygote | heterozygote | NM_000441(*SLC26A4*):c.1983C>A(p.D661E) | heterozygote | LP |

| Sample ID | ECMA results | | Sanger sequencing results | | | Pathogenicity classification |
|---|---|---|---|---|---|---|
| | Locus | Genotypic status | Genotypic status of ECMA locus | Valid Locus | Genotypic status of valid Locus | |
| 463 | NM_000527(*LDLR*):c.1197_1205del(p.399_402del) | heterozygote | wild type | NM_000527(*LDLR*):c.1186G>A(p.Gly396Ser) | heterozygote | LP |
| 465 | NM_000137(*FAH*):c.1056C>A(p.S352R) | homozygote | heterozygote | NM_000137(*FAH*):c.1056C>T(p.Cys352=) | heterozygote | B |
| 474 | NM_023110(*FGFR1*):c.1854G>T(p.K618N) | heterozygote | wild type | - | wild type | - |
| 483 | NM_001844(*COL2A1*):c.1648C>T(p.R550C) | heterozygote | heterozygote | NM_001844(*COL2A1*):c.1648C>T(p.R550C) | heterozygote | VUS |
| 483 | NM_000169(*GLA*):c.128G>T(p.G43V) | homozygote | wild type | - | wild type | - |
| 484 | NM_000152(*GAA*):c.1580_1581del(p.R527fs) | homozygote | wild type | - | wild type | - |
| 485 | NM_000053(*ATP7B*):c.2333G>T(p.R778L) | heterozygote | heterozygote | NM_000053(*ATP7B*):c.2333G>T(p.R778L) | heterozygote | P |
| 485 | NM_000053(*ATP7B*):c.2309T>C(p.L770P) | heterozygote | wild type | NM_000053(*ATP7B*):c.2310C>G(p.Leu770=) | heterozygote | VUS |
| 490 | NM_000152(*GAA*):c.1580_1581del(p.R527fs) | homozygote | wild type | - | wild type | - |
| 492 | NM_000033(*ABCD1*):c.1537A>C(p.K513Q) | homozygote | wild type | - | wild type | - |
| 496 | NM_001005741(*GBA*):c.1448T>C(p.L483P) | heterozygote | wild type | - | wild type | - |
| 497 | NM_000277(*PAH*):c.157C>T(p.R53C) | heterozygote | wild type | NM_000277(*PAH*):c.158G>A(p.Arg53His) | neighboring mutati | VUS |
| 501 | NM_001005741(*GBA*):c.928A>G(p.S310G) | heterozygote | wild type | - | wild type | - |
| 501 | NM_001005741(*GBA*):c.721G>A(p.G241R) | heterozygote | wild type | - | wild type | - |
| 508 | NM_000543(*SMPD1*):c.839_840insCATCCCCG(p.D28 | heterozygote | heterozygote | NM_000543(*SMPD1*):c.839_840insCATCCCCG(p.D280fs) | heterozygote | P |
| 508 | NM_000543(*SMPD1*):c.1458T>G(p.S486R) | heterozygote | heterozygote | NM_000543(*SMPD1*):c.1458T>G(p.S486R) | heterozygote | P |
| 509 | NM_000387(*SLC25A20*):chr3：48884134A>C | homozygote | homozygote | NM_000387(*SLC25A20*):c.199-10T>G | homozygote | P |
| 510 | NM_000053(*ATP7B*):c.3809A>G(p.N1270S) | heterozygote | heterozygote | NM_000053(*ATP7B*):c.3809A>G(p.N1270S) | heterozygote | P |
| 510 | NM_000053(*ATP7B*):c.1708-1G>C | heterozygote | heterozygote | NM_000053(*ATP7B*):c.1708-1G>C | heterozygote | P |
| 511 | NM_000053(*ATP7B*):c.2804C>T(p.T935M) | heterozygote | heterozygote | NM_000053(*ATP7B*):c.2804C>T(p.T935M) | heterozygote | P |
| 511 | NM_000053(*ATP7B*):c.2075T>C(p.L692P) | heterozygote | heterozygote | NM_000053(*ATP7B*):c.2075T>C(p.L692P) | heterozygote | LP |
| 512 | NM_000142.5(*FGFR3*):c.1138G>A(p.G380R) | heterozygote | heterozygote | NM_000142.5(*FGFR3*):c.1138G>A(p.G380R) | heterozygote | P |

ECMA: expanded chromosomal microarray; P: pathogenic; LP: like pathogenic; VUS: variant of uncertain significance; LB: likely benign; B: benign.

**Table S7. Clinical indications and pregnancy outcomes of 13 fetuses with pathogenic or likely pathogenic SNVs detected by ECMA**

| sample ID | Clinical indications | Pregnancy outcomes | Single nucleotide polymorphisms | Variant classification | Genotypic status | ECMA results | Sanger sequencing results |
|---|---|---|---|---|---|---|---|
| 46 | Shortened emur, humerus, and tibia | abortion | NM_000142(*FGFR3*):c.1138G>A(p.G380R) | P | heterozygote | 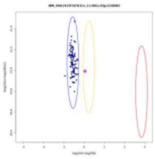 | 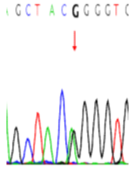 |
| 246 | Mother with a history of 3 miscarriages | declined follow-up | NM_000441(*SLC26A4*):c.697G>C(p.V233L) | LP | heterozygote | 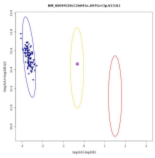 | 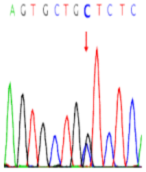 |
| | | | NM_000441(*SLC26A4*):c.680C>G(p.Ala227Gly) | LP | heterozygote | 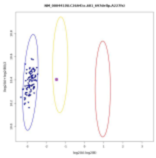 | 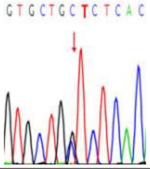 |
| 322 | Left superior vena cava with coronary sinus dilation | liveborn | NM_000518.5(*HBB*): c.25_26insG(p.Lys9ArgfsTer15) | LP | heterozygote | 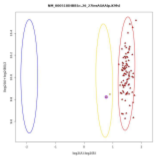 | 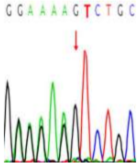 |
| 323 | Small double apex diameter | liveborn | NM_000518.5(*HBB*): c.25_26insG(p.Lys9ArgfsTer15) | LP | heterozygote | 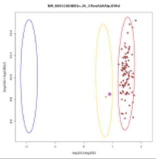 | 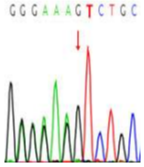 |
| 327 | Mother with a history of 2 miscarriages | declined follow-up | NM_000089(*COL1A2*):c.3794C>G(p.S1265C) | LP | heterozygote | 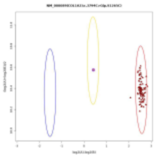 | 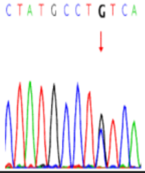 |
| 341 | Tetralogy of Fallot | declined follow-up | NM_000531(*OTC*):c.419C>T(p.Ala140Val) | LP | heterozygote | 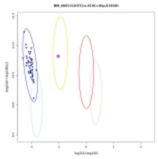 | 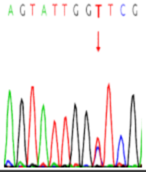 |

| sample ID | Clinical indications | Pregnancy outcomes | Single nucleotide polymorphisms | Variant classification | Genotypic status | ECMA results | Sanger sequencing results |
|---|---|---|---|---|---|---|---|
| 444 | Mother older than 35 | liveborn | NM_000384(*APOB*): c.10579C>T(p.R3527W) | LP | heterozygote | 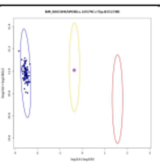 | 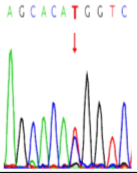 |
| 463 | Mother with chromosome translocation | declined follow-up | NM_000527(*LDLR*):c.1186G>A(p.Gly396Ser) | LP | heterozygote | 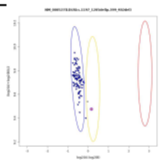 | 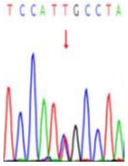 |
| 508 | Sibling with an *SMPD1* variants passed away | abortion | NM_000543(*SMPD1*):c.839_840insCATCCCCG(p.D280fs) | P | heterozygote | 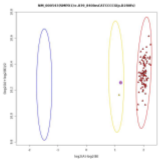 | 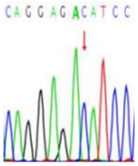 |
| | | | NM_000543(SMPD1):c.1458T>G(p.S486R) | P | heterozygote | 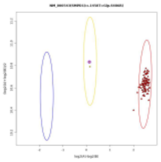 | 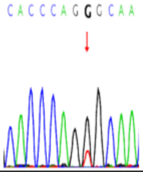 |
| 509 | Parents are consanguineously married | abortion | NM_000387.6(*SLC25A20*):c.199-10T>G | LP | homozygote | 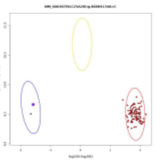 | 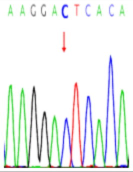 |
| 510 | Sibling with Wilson's Disease | liveborn | NM_000053(*ATP7B*): c.3809A>G(p.N1270S) | P | heterozygote | 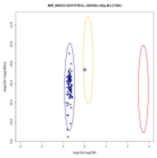 | 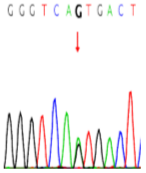 |
| | | | NM_000053(*ATP7B*): c.1708-1G>C | P | heterozygote | 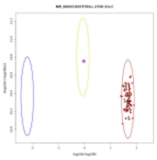 | 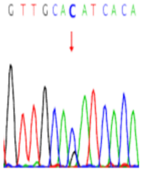 |

| sample ID | Clinical indications | Pregnancy outcomes | Single nucleotide polymorphisms | Variant classification | Genotypic status | ECMA results | Sanger sequencing results |
|---|---|---|---|---|---|---|---|
| 511 | Widened bilateral renal pelvis | liveborn | NM_000053(*ATP7B*): c.2804C>T(p.T935M) | P | heterozygote | 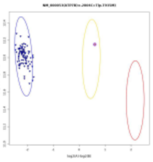 | 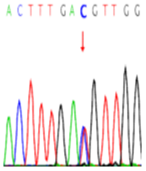  |
| | | | NM_000053(*ATP7B*): c.2075T>C(p.L692P) | LP | heterozygote | 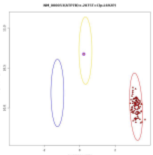 | 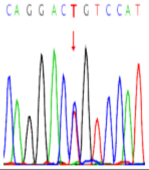  |
| 512 | Nuchal translucency =3.1mm | abortion | NM_000142.5(*FGFR3*):c.1138G>A(p.G380R) | P | heterozygote | 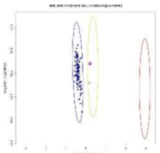 | 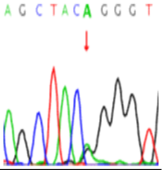  |

SNVs: single nucleotide variants; ECMA: expanded chromosomal microarray; P: pathogenic; LP: like pathogenic.
